# From feasibility to neuroanatomic validity of remote cognitive smartphone assessments in early Alzheimer’s disease

**DOI:** 10.64898/2026.05.19.26353554

**Authors:** Kirsten I. Taylor, Arnaud M. Wolfer, Irma T. Kurniawan, Miguel Veloso, Goullou Keita, Niels Hagenbuch, Beijue Shi, Foteini Orfaniotou, Eduardo A. Aponte, Macarena Garcia Valdecasas Colell, Christopher H. Chatham, Štefan Holiga, Raphael Ullmann, Wael Abouelkheir, Simone Rey-Riek, Emma Poon, David Watson, Mercè Boada, Thanneer M. Perumal

## Abstract

Digital health technologies (DHT) offer a promising solution to the timely identification of early Alzheimer’s disease (eAD) to enable early treatment. This study evaluated the feasibility, acceptability, adherence, reliability, and preliminary clinical and content validity of the novel AD Digital Assessment Suite (AD-DAS). 123 individuals (32 healthy controls (HC), 31 amyloid-PET negative (SCDn), 30 amyloid-PET positive (SCDp) with subjective cognitive decline, and 30 early AD (eAD)) participated. AD-DAS was remotely deployed for 28 days. Remote testing was feasible (97.6% completers), acceptable (>85% “good”), and associated with high adherence (96%). Metrics showed moderate to excellent test-retest reliability (ICC 0.53–0.91), associations with clinical comparators (adjusted R^2^ 0.01-0.24), differentiated eAD from other known groups (absolute log odds differences 0.6–3.28), and correlated with brain atrophy in expected regions. Episodic and working memory AD-DAS metrics differentiated SCDp from SCDn participants. These preliminary findings suggest that AD-DAS may be a promising tool for detecting cognitive impairments in early AD stages.

## Introduction

The rising prevalence of Alzheimer’s disease (AD)^1^ and the advent of disease-modifying therapies are converging to build an urgent need to efficiently and accurately identify individuals in the earliest stages of AD who may benefit from such interventions^2,3^. Digital health technology (DHT) tools such as smartphone-based cognition applications are proposed as one solution to this challenge: remote testing would reduce the need for time-intensive administration of cognitive tests in the clinical setting by well-trained psychometrists, thereby reducing burden on healthcare resources; cognitive tests could be frequently administered to generate more reliable estimates of performance^4^; and the tasks could be deployed very broadly (i.e. any smartphone owner)^5,6^. However, key open questions remain regarding the appropriate design of such a DHT testing solution, the feasibility and acceptability of self-administered cognitive testing on smartphones at home, and the ensuing reliability, validity and sensitivity of their outcome measures.

The earliest detection of AD typically occurs in a primary care setting. Several multi-domain, very short (ca. 10 minutes) and short (ca. 20 minutes) case-finding tools have been proposed for this setting, whereby, as expected, the sensitivity and specificity for dementia detection increases with examination duration^7^. Several concerns have been raised regarding current case-finding procedures and tools to detect preclinical and early AD. A key limitation is the burden to the healthcare systems, including training requirements and administration time, which is considered high even with very short instruments^8^. Indeed, these restrictions translate into the very brief instruments being recommended to minimize training and administration times, with ramifications for diagnostic accuracy in dementia and misdiagnosis of premorbid cognitive impairment and delirium^8^. An additional driver of the predictive accuracy appears to be the robustness of the training protocols, as indicated by the proportion of administration and scoring errors (i.e. 23%) across a variety of case-finding instruments used in the primary care setting^9^. Notably, this error rate is comparable to that found with raters trained to conduct screening for clinical trials (i.e., 24% for MMSE)^10^. Moreover, brief case-finding tools are relatively insensitive to the prodromal stages of AD^8,11,12^, i.e., those stages when disease-modifying therapy is hypothesized to be most impactful^13^. One key reason for this may be that the earliest cognitive manifestation of incipient AD – decline in long-term episodic memory functioning^12^ – cannot be adequately assessed in very short testing sessions that necessarily limit the length of the delayed recall period of the administered memory test. Finally, an often under-discussed component of efficient detection of preclinical and early AD is the importance of differential diagnostic measures. Specifically, case-finding tools ideally provide information about potential non-AD causes of cognitive impairment^14^, most notably reversible causes of cognitive impairment including depression, as well as vascular dementia^15^. This would require the assessment of a broader range of cognitive domains than those affected in typical AD.

DHTs may represent one method to meet these challenges^14^. Cognitive tests can be self-administered at home, e.g., on smartphones, enabling a more in-depth assessment of a given cognitive function (e.g., long-term memory over days) and the testing of a broader range of cognitive functions to support differential diagnostic considerations. Moreover, the ability to test cognitive performance on multiple days reduces potentially confounding effects such as sleep and mood on cognitive performance^16^, thereby generating more reliable outcome metrics^6^.

Different remote cognitive testing solutions have been proposed, varying from individual memory tests^6,17^ to those aiming to provide a broader assessment of cognitive functioning^14^. An example of the latter is the Cogstate Brief Battery (CBB), which comprises tests of processing speed, visual attention, visual learning, and working memory. The feasibility and utility of remote, web-based CBB administration on individuals’ private laptops or desktops were found to be high in a non-demented, community-based sample with a family history of dementia^18^. In a substudy of ADNI-2, baseline completion of the CBB at home was shown to be feasible, at-home CBB metrics were found to be concordant with in-clinic CBB metrics, and these metrics differentiated healthy controls from Mild Cognitive Impairment (MCI) and AD groups^19^. An alternative approach to remote cognitive testing is to provide very brief (i.e., several minutes) Ecological Momentary Assessments (EMAs) comprising different ultra-short cognitive tests multiple times throughout the day^20^. Two apps have pioneered this approach: (1) the Mobile Monitoring of Cognitive Change (M2C2)^4,21^ measuring processing speed, visual short-term memory binding, and visuospatial working memory, and (2) the Ambulatory Research in Cognition (ARC) app measuring processing speed, associative memory, and visuospatial working memory^22^. The EMA approach was found to be feasible and acceptable to participants, provided robust aggregate test measures with high reliability, correlated with standard neuropsychological tests and, in individuals at risk of developing AD, with AD biomarkers^4,22^. These findings indicate that remote cognitive testing approaches may overcome current and future hurdles faced by the healthcare system in identifying individuals on the AD spectrum.

The purpose of the present study was to investigate a novel smartphone DHT for remote cognitive testing – the AD Digital Assessment Suite (AD-DAS). The AD-DAS was designed to meet the following two primary goals: (1) to robustly assess cognitive functions known to be impacted in preclinical and early AD, in particular, episodic memory with long-term delays over days, and (2) to measure other key cognitive and motor domains for differential diagnostic information and potential utility beyond AD. The present study aimed to quantify the feasibility, acceptability of, and adherence to repeated, at-home cognitive testing, and the preliminary reliability and validity of the DHT-derived cognitive outcome metrics. Three aspects of validity of the DHT-derived outcome measures were assessed: (1) relationships with standard in-clinic neuropsychological tests, (2) ability to discriminate between known groups, and (3) voxel-based morphometry (VBM) analyses of anatomic magnetic resonance imaging (MRI) scans which related DHT performance to regional atrophy estimates across the entire brain. These VBM results offer an orthogonal set of data with which to assess the validity of the DHT cognitive outcome metrics, i.e., whether they were associated with atrophy in the expected, but not in the unexpected, brain regions. Finally, to estimate the ability of the AD-DAS tasks to detect cognitive changes associated with preclinical AD, the present study included individuals with Subjective Cognitive Decline (SCD), i.e., those who perceive that their cognition has declined from a previous level of functioning, yet do not exhibit cognitive impairment on formal clinical neuropsychological testing^23^. While SCD is associated with an increased risk of developing mild cognitive impairment (MCI) and dementia within a five-year timeframe (HR 1.9 and HR 1.7, respectively)^24^, the majority of SCD individuals do not progress to these syndromes within several years^25^. For this reason, the present study included both SCD individuals who were amyloid PET negative (SCDn) and positive (SCDp), as the latter are known to exhibit an increased rate of future cognitive decline presumed to be associated with AD pathology^26^. Thus, performance in the SCDn and SCDp groups were compared to explore the ability of the AD-DAS to detect subtle cognitive changes associated with preclinical AD.

## Methods

### Study Design

#### Population

A total of 123 individuals participated across 3 sites in the United States and 2 sites in Spain. The cohort included 32 amyloid-negative healthy volunteers (HC), 61 individuals with subjective cognitive decline (SCD), 31 of whom were amyloid-negative (SCDn) and 30 were amyloid-positive (SCDp), and 30 individuals with early AD (eAD; all with confirmed amyloid PET positivity). All participants were 65 years of age or older, had prior experience with smartphone or tablet technology, and had normal or corrected-to-normal visual and auditory acuity. They were fluent in the study languages of English or Spanish (as per the judgment of the investigator), willing and able to undergo magnetic resonance imaging (MRI) scanning, and had a study partner (informant) willing to participate throughout the study. Importantly, none of the participants had a history or known presence of any neurological or psychiatric disorder (beyond the diagnosis required for study inclusion), or were taking any medication known to affect cognitive functioning. None of the HCs or SCD participants had a prior diagnosis and/or treatment for a memory disorder. HC participants’ neuropsychological test scores in the Free and Cued Selective Reminding Test (FCSRT)^27^, Trail-Making Test (TMT)^28^ Parts A&B, Category Fluency Test (Animals) (VFT)^29^, Digit Symbol Coding Test (DSCT)^30^ and Boston Naming Test (BNT)^31^ (see below) were within one standard deviation (SD) of mean normative values, and all achieved a Cognitive Change Index (CCI)^32^ score of < 16 on the first 12 questions^33^. SCD participants were included if the CCI score was ≥ 16 based on the first 12 questions, the Mini-Mental State Examination (MMSE)^34^ score ≥ 27 for participants with at least a high school graduation and otherwise ≥ 26^35^ and if the Clinical Dementia Rating (CDR)^36^ global score was 0. eAD participants were diagnosed with MCI due to AD or probable AD as per the NIA-AA criteria^37^. All had an MMSE score ≥ 24 for those with at least a high school graduation and otherwise ≥ 23 points, and a CDR global score of 0.5.

The demographic and clinical characteristics of all recruited participants are listed in Table 1. Among the 123 recruited participants, the majority were White (94.3%), not Hispanic or Latino (69.1%) and male (57.7%); the median age at baseline was 71 years (range: 65−89). Groups were comparable with respect to age (F(3) = 2.08; p = 0.11), gender distribution (χ2(3) = 1.017, p = 0.80), ethnicity (χ2(3) = 4.75, p = 0.19), or educational attainment (χ2(3) = 8.67, p = 0.47).

**Table 1.** Demographic and clinical characteristics of study participants (mean (standard deviation), [range]).

|  | HC | SCDn | SCDp | eAD |
| --- | --- | --- | --- | --- |
| n | 32 | 31 | 30 | 30 |
| Age (y) | 70.3 (5.1)<br>[65 – 83] | 70.9 (5.3)<br>[65 - 89] | 71.8 (5.3)<br>[65 - 82] | 73.9 (5.2)<br>[65 - 83] |
| Sex (M:F) | 20:12 | 17:14 | 16:14 | 18:12 |
| Ethnicity (Hispanic or Latino - yes:no) | 14:18 | 6:25 | 10:20 | 8:22 |
| Educational attainment (elementary school:some high school: high school: some college: college graduate: post-graduate) | 4:7:7:2:6:6 | 2:1:7:5:10:6 | 4:4:6:1:7:8 | 6:4:3:3:8:6 |
| Country (USA:Spain) | 17:15 | 17:14 | 14:16 | 16:14 |
| MMSE total score | 29.3 (0.5)<br>[29 - 30] | 29.1 (0.8)<br>[27 - 30] | 29 (1.1)<br>[27 - 30] | 26 (1.8)<br>[23 - 30] |
| CCI self - report | 13.3 (1.2)<br>[12 - 15] | 22.4 (4.9)<br>[16 - 36] | 23.5 (5.8)<br>[16 - 43] | 28.2 (6.8)<br>[14 - 42] |
| CCI informant report | 22.1 (3.5)<br>[20 - 36] | 26.6 (6)<br>[20 - 48] | 29.1 (9.4)<br>[20 - 62] | 45.7 (13.7)<br>[23 - 80] |
| A - IADL - Q - SV Total Score | 68.0 (3.1)<br>[58.2 - 70] | 68.3 (2.7)<br>[58.9 - 70] | 67.4 (3.8)<br>[51.7 - 69.9] | 61.2 (7.9)<br>[41.3 - 69.8] |
| FCSRT Free Recall (Encoding) | 36.9 (4)<br>[30 - 44] | 32.2 (6)<br>[21 - 44] | 34.6 (5.4)<br>[25 - 48] | 22.7 (9.8)<br>[0 - 37] |
| FCSRT Sensitivity to Cueing (Encoding) | 1.0 (0.3)<br>[0.8 - 2.7] | 1.0 (0.1)<br>[0.9 - 1.6] | 1.0 (0.0)<br>[0.9 - 1.0] | 0.9 (0.2)<br>[0.4 - 1.0] |
| FCSRT Free Recall (30 min Delay) | 12.6 (1.8) | 10.8 (2.2) | 12.6 (1.8) | 7.0 (4.3) |
|  | [9.0 - 16.0] | [6.0 - 15.0] | [8.0 - 16.0] | [0.0 - 12.0] |
| TMT Time Taken Part A | 17.9 (0.9)<br>[16.0 - 19.0] | 43.0 (15.9)<br>[19.0 - 85.0] | 37.3 (13.8)<br>[17.4 - 79.0] | 64.2 (31.9)<br>[27.0 - 147.0] |
| TMT Time Taken Part B - Time<br>Taken Part A | 21.2 (1.4)<br>[18.0 - 24.0] | 53.1 (35.6)<br>[5.0 - 164.0] | 57.5 (68.8)<br>[12.0 - 397.0] | 92.3 (60.0)<br>[18.0 - 281.0] |
| BNT Correct Responses | 53.6 (17.6)<br>[0 - 60] | 55.7 (4.9)<br>[43 - 60] | 55.9 (11)<br>[0 - 60] | 50.8 (12.5)<br>[0 - 60] |
| VFT Ratio Acceptable Words to<br>Words Generated | 1.0 (0.0)<br>[0.9 - 1.0] | 1.0 (0.0)<br>[0.9 - 1.0] | 1.0 (0.0)<br>[0.9 - 1.0] | 0.9 (0.2)<br>[0.2 - 1.0] |
| DSCT Number of Correct Answers | 51.3 (16.8)<br>[0 - 89] | 48.2 (10.1)<br>[27 - 66] | 47.5 (11.7)<br>[29 - 70] | 36.4 (15.6)<br>[13 - 75] |
| T25FWT Average time taken<br>[seconds] | 10.5 (3.88)<br>[4.5 - 22] | 8.74 (3.62)<br>[4.6 - 17] | 10.59 (3.66)<br>[4 - 16.5] | 13.12 (5.94)<br>[4 - 27.5] |
| TUG Time take to complete task<br>[seconds] | 7.84 (3.22)<br>[4 - 20] | 9.6 (3.61)<br>[4 - 20] | 8.53 (2.97)<br>[5 - 18] | 10.86 (5.14)<br>[5 - 30] |
| HADS Anxiety | 2.3 (3)<br>[0 - 12] | 3.3 (3.7)<br>[0 - 17] | 3.2 (3.1)<br>[0 - 11] | 3.9 (3.7)<br>[0 - 15] |
| HADS Depression | 1.7 (2.8)<br>[0 - 8] | 1.7 (1.8)<br>[0 - 6] | 2.1 (2.4)<br>[0 - 8] | 2.6 (2.8)<br>[0 - 9] |
| PSQI Global score | 3 (3)<br>[0 - 13] | 3.8 (2.4)<br>[0 - 9] | 3.9 (2.7)<br>[0 - 10] | 4.5 (3.7)<br>[1 - 13] |
| NPI Total Score | 0.9 (1.7)<br>[0 - 6] | 1 (2.5)<br>[0 - 12] | 1.3 (2.9)<br>[0 - 11] | 1.4 (1.8)<br>[0 - 6] |
| UCLA Loneliness Scale Total Score | 44.6 (7.6)<br>[30 - 62] | 41 (10.2)<br>[23 - 56] | 43.5 (9)<br>[23 - 59] | 42 (10.2)<br>[20 - 57] |
| SNI Total Number of people in social<br>network | 16.6 (8.9)<br>[6 - 46] | 14.3 (9.7)<br>[3 - 40] | 13.8 (6.8)<br>[4 - 31] | 11.7 (5.3)<br>[1 - 23] |

We note that SCDn participants, on average, had higher education than other groups. Consistent with the inclusion/exclusion criteria, MMSE scores indicated greater impairment in the eAD relative to all other groups, and CCI informant and self-report scores were greater in the SCD and eAD compared to the HC groups. The difference between informant and self-report CCI scores was much larger for the eAD group compared to all other groups. Mean group scores on the clinical scales were consistent with the inclusion/exclusion criteria and are in the expected directions.

This study was conducted in full conformance with the ICH E6 guideline for Good Clinical Practice and the principles of the Declaration of Helsinki, or the laws and regulations of the country in which the research is conducted, whichever affords the greater protection to the individual. Ethics committees at each site approved the study: CEIC Hospital de Bellvitge (approval number PR455/20), Stanford University Institutional Review Board (approval number 55763), CEIC Fundacion Jimenez Diaz, Area Administrativa de Investigacion (approval number 21/20), WIRB-Copernicus Group (WCG) USA (approval number 20202969) and Advarra USA (approval number Pro00046964).

### Procedures

The study consisted of a screening period of up to 12 weeks, followed by a baseline in-clinic visit, 30-day remote monitoring period, and in-clinic end-of-study visit.

#### Screening

The screening visit(s) entailed history-taking, a general physical examination, administration of clinical and neuropsychological assessments, including the CDR Scale^36^, MMSE^34^, CCI^38^, FCSRT^27^, TMT^28^, VFT^29^, DSCT^30^, BNT^31^, Amsterdam instrumental activities of daily living questionnaire short version (A-IADL-Q-SV)^39^, required for inclusion/exclusion criteria. Eligibility was confirmed by MRI and amyloid PET scan, both read centrally.

#### Baseline (Day 1)

At the baseline in-clinic study visit, eligible participants received a locked-down, pre-configured study smartphone (Galaxy A6, Samsung, Seoul, South Korea) for daily remote self-assessments and training for performing smartphone tasks at home. Instructions for each task were presented prior to each task administration throughout the study. The study team stressed the importance of the participants completing the tasks on their own, i.e., without help from others, including the study partner. All participants were instructed to complete a pre-configured set of active tasks at the same time each day, ideally in the morning, and to recharge the smartphones overnight. Additionally, vital signs and a general medical assessment, the Timed 25-foot Walk Test (T25WT^40^) and the Timed Up and Go test (TUG^41^) were administered at the baseline visit.

#### Remote Monitoring (Days 2–31)

Participants completed a subset of AD-DAS tasks remotely during days 2–31, with a two-day break in between, and according to a pre-programmed schedule of assessments on the smartphone. Each task was scheduled for administration on at least 9 different days over the course of 28 days of remote testing. App data were encrypted and uploaded to secure servers when the smartphones were connected to WiFi; at a minimum, this occurred during the site visits.

#### End of study (Day 32)

Participants returned their study smartphones at an end-of-study in-clinic visit on day 32, and reported their experience regarding smartphone use via the User Experience Survey (see Supplementary material). In addition, the study partners provided information on the support they provided to the participants during the remote monitoring phase (Study Partner Survey; see Supplementary material). Final assessments include vital signs and a medication check.

### Assessments

#### Clinical and Neuropsychological Scales

All clinical and neuropsychological scales administered during screening and baseline visits are described in the ‘Study Design - Population’ sub-section of Methods in and were administered according to standard procedures.

#### Amyloid PET Imaging

A centrally read amyloid PET imaging scan was required for all participants to confirm their amyloid positivity/negativity status. Historical amyloid PET scans were accepted if they had been acquired within six months prior to the signing of the informed consent form. If a new PET scan was necessary to meet the inclusion/exclusion criteria, one of the following radioligands was used: ^18^F florbetapir (Amyvid™), ^18^F flutemetamol (Vizamyl™), or ^18^F florbetaben (Neuraceq™).

#### Anatomic MRI Imaging

All images were acquired with comparable 3T MRI scanners using standard structural T1 sequences (voxel resolution of 1×1×1.2 mm) harmonized across all five sites by a centralized imaging vendor. Two study sites used a 3D[MPRAGE sequence on Siemens MRI scanners, two sites used a 3D-SPGR sequence on a GE MRI scanner, and one site used a 3D T1TFE on a Philips MRI scanner.

#### Alzheimer’s Disease Digital Assessment Suite (AD-DAS)

The AD-DAS comprises nine active tasks described in Table 2 and described below. Three active tasks (i.e., Gallery Game, Story Time and Tilt Task) were developed in a collaboration between the Oxford Institute of Digital Health, Roche and Eli Lilly.

**Table 2.**
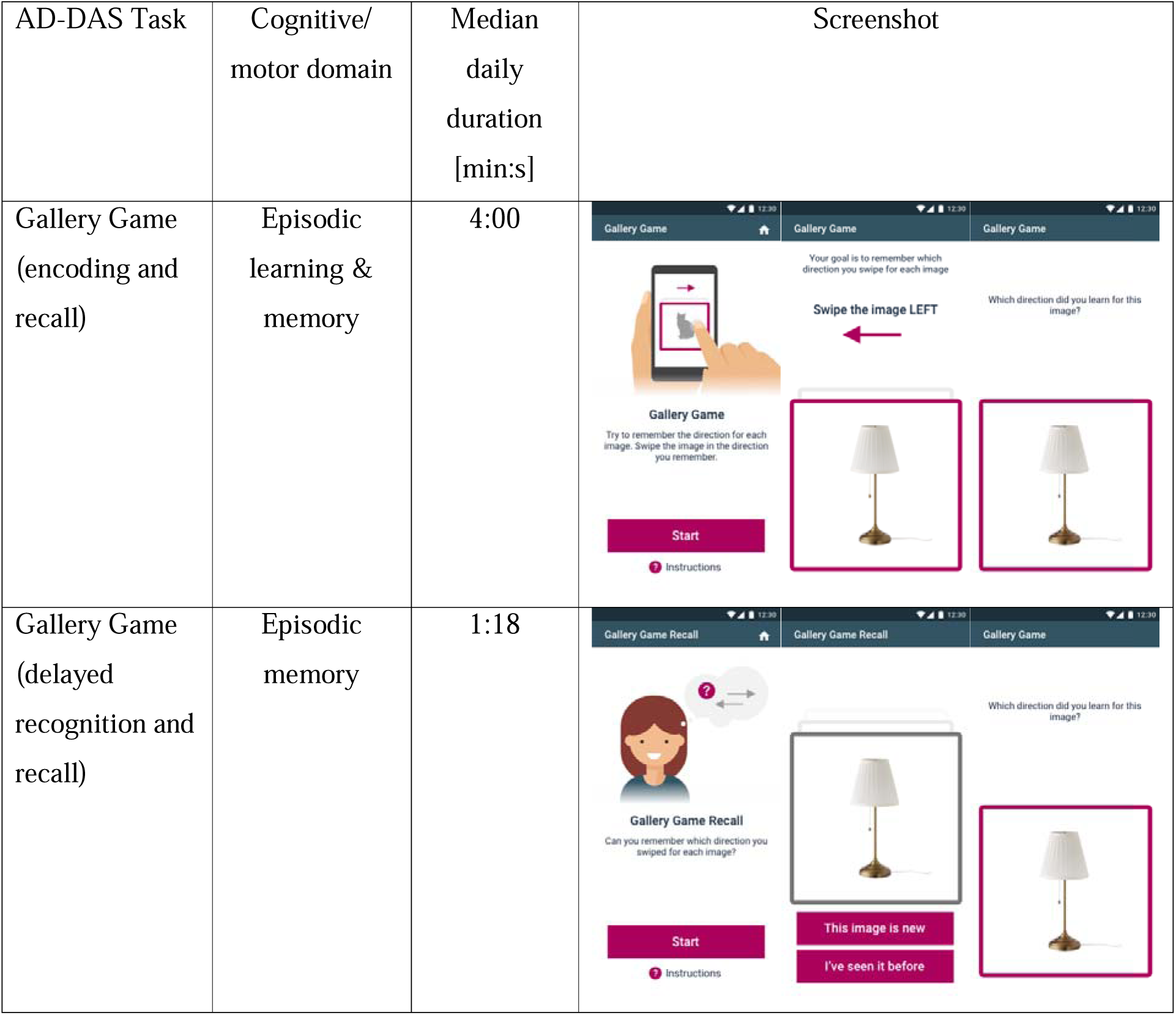
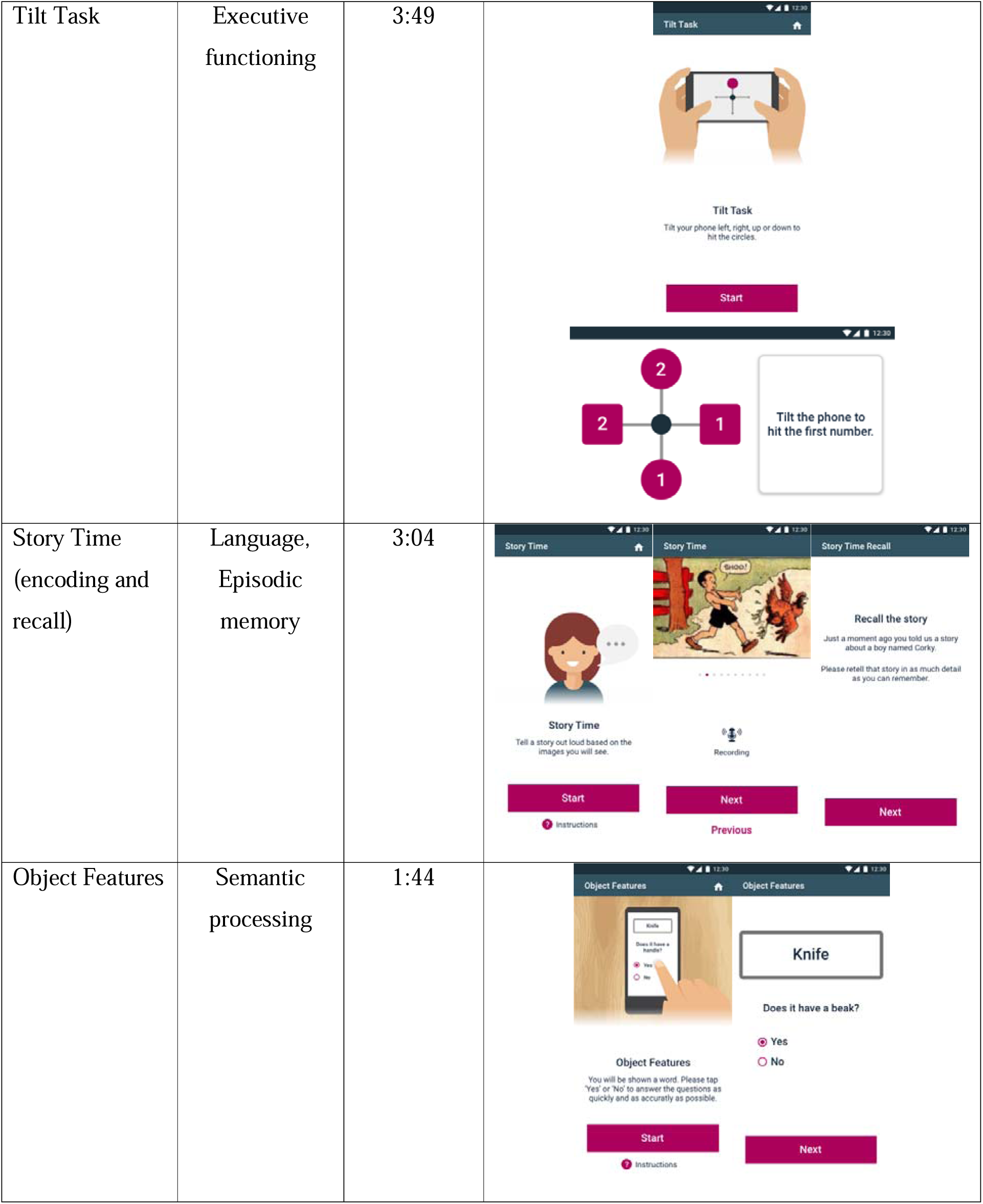
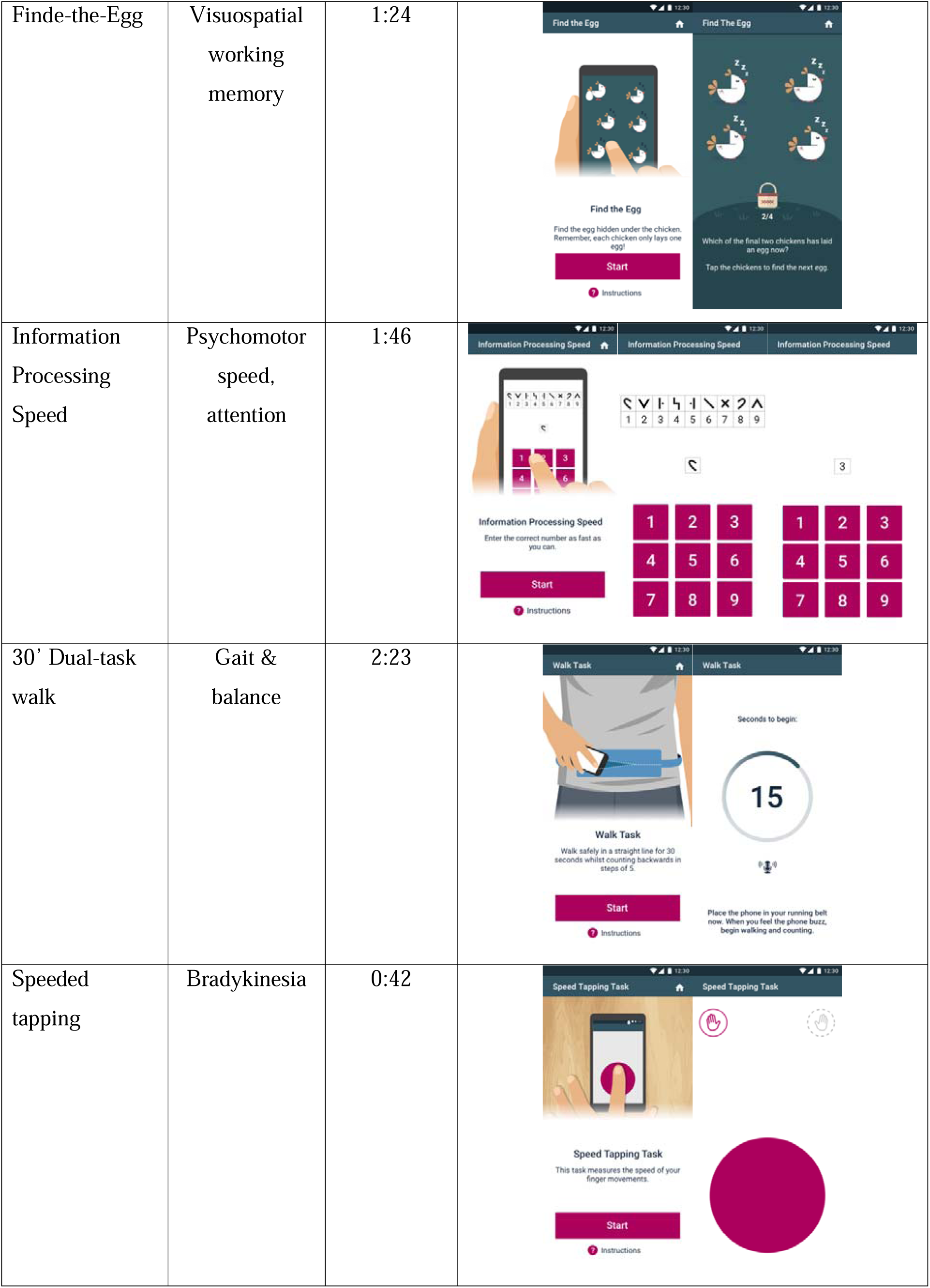
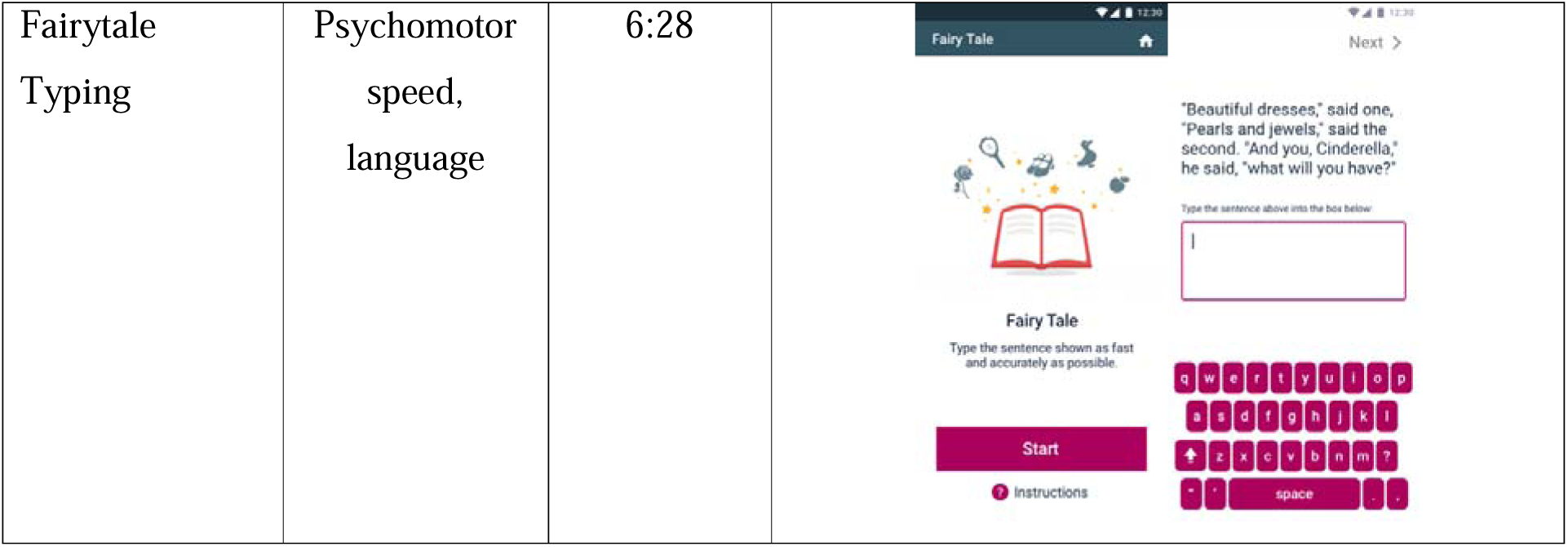
AD-DAS task names, cognitive/motor domains, median daily duration, and selected screenshots.

| AD-DAS Task | Cognitive/<br>motor domain | Median<br>daily<br>duration<br>[min:s] | Screenshot |
| --- | --- | --- | --- |
| Gallery Game<br>(encoding and<br>recall) | Episodic<br>learning &<br>memory | 4:00 |  |
| Gallery Game<br>(delayed<br>recognition and<br>recall) | Episodic<br>memory | 1:18 |  |
| Tilt Task | Executive functioning | 3:49 |  |
| Story Time (encoding and recall) | Language, Episodic memory | 3:04 |  |
| Object Features | Semantic processing | 1:44 |  |
| Finde-the-Egg | Visuospatial<br>working<br>memory | 1:24 |  |
| Information<br>Processing<br>Speed | Psychomotor<br>speed,<br>attention | 1:46 |  |
| 30' Dual-task<br>walk | Gait &<br>balance | 2:23 |  |
| Speeded<br>tapping | Bradykinesia | 0:42 |  |
| Fairytale<br>Typing | Psychomotor<br>speed,<br>language | 6:28 |  |

#### Gallery Game

The Gallery Game was designed to assess episodic learning and memory and has three components: learning (Gallery Game Learning), delayed object recognition (Gallery Game Recognition) and free recall (Gallery Game Free Recall)^42^. Briefly, participants were instructed to learn which direction specific photorealistic pictured objects belonged with: left, right, or up. Learning trials presented participants with the single objects with and without arrows, and participants were instructed to learn which direction an object is associated with and to swipe the object off the screen in the corresponding direction. Participants started with a stack of 2 objects, each with an arrow. Objects were increasingly added to the stack each time the preceding object-swipe direction associations had been learned, as defined by correct direction recall to images presented without an arrow. Throughout, the order of objects presentation was pseudorandomized with the constraint that the same object was not shown in succession. The learning task ended when 6 object-arrow pairs had been added to the stack and each object-swipe direction had been correctly recalled on at least 5 of the last 6 presentations. Either during the same session (i.e., following a delay period varying by day in the study: 6 x 5 minutes, 3 x 15 minutes, 3 x 30 minutes, and 6 x 60 minutes delay) or later (i.e., 2 x 1 day, 1 x 3 days, 1 x 6 days), the Gallery Game Recognition task showed participants a stack of photorealistic pictured objects, 50% of which had been shown during learning (targets) and 50% of which were new (distractors). Participants were instructed to decide whether each object was old or new and to select the corresponding button (“I’ve seen it before” vs. “This image is new”, respectively). Following Recognition, the Gallery Game Free Recall task presented participants with the target objects without arrows, three trials for each object, and instructed participants to select a button corresponding to the direction the object had been associated with. The majority response was defined as the Free Recall response for the given object.

Gallery Game Learning performance was quantified with the proportion of correct swipes for objects shown without arrows, and the total time to complete the task. Gallery Game Recognition performance was quantified with total time taken to complete the task (as a proxy of difficulty performing the task) and discriminability, i.e. balanced accuracy defined as the ratio of correctly identified targets and distractors to the total number of objects presented.

Gallery Game Free Recall performance was quantified with total time to complete the task and the average percentage of correctly identified object-swipe direction associations.

Outcome measures for both memory tasks were calculated separately for tasks conducted within 24 hours of learning and those conducted at least 24 hours after learning.

#### Tilt Task

The Tilt Task was designed to assess executive functioning and response inhibition based on principles of the Trail-Making Test^28^ and Stroop interference test^43^, respectively. This task instructed participants to hold the smartphone in landscape mode, and to tilt the smartphone to move a ball at the center of a cross on the screen either up, down, right or left to ‘hit’ target stimuli located at the ends of the cross according to an increasingly complex set of rules (corresponding to ‘levels’ of the task): (1) if the target stimulus was a filled circle (versus no stimulus); (2) to hit ascending numbers presented in squares but not numbers presented in circles; (3) same as (2) but during half of the trials, the central black circle changed to a star, at which time participants were required to tilt the phone in the *opposite* direction to hit the intended target (‘starred trials’); (4) increasing numbers when squares were shown, and increasing letters when circles were shown, with both numbers and letters shown on all trials; (5) same as (4) with starred trials. Level (1) presented 16, and all other levels 32 randomized stimuli. If three errors were made during any level, the task stopped, and participants were provided with an option to redo the level. Only responses from the first administration of each level were analyzed.

Tilt Task performance was quantified with a total score metric defined as the highest level successfully completed.

#### Story Time

Story Time was designed to assess language fluency and content through two tasks: Story Time Narration, where participants told a story out loud based on nine-panel cartoon strips, and Story Time free recall, which evaluated episodic memory through the recall of the story, similar to the Logical Memory task^44^. Participants viewed one panel of the cartoon strip at a time, navigating the screens at their own pace using “NEXT” and “PREVIOUS” buttons while narrating the story.

Following the narration, Story Time free recall sessions (i.e., recalling the story in absence of the stimuli) took place (a) immediately after narration (Story Time Immediate Free Recall) and (b) after a delay greater than 24 hours (Story Time Delayed Free Recall). During both recall sessions, participants were instructed to retell as much of the story as they could remember without viewing the cartoon strip.

Speech was recorded using the smartphone’s microphone. Some participants were exposed to multiple stories during the learning phase as a result of not having completed the task in a single session within a given calendar day. As a result, a few instances of delayed recall sessions involved the re-telling of multiple stories. The affected sessions were manually excluded from the analysis during data quality control.

Speech data were processed through an automated speech segmentation and a manual transcription pipeline (see Supplementary Material for processing details), generating the duration of active speech and pauses, and noun rate (i.e., number of nouns divided by the total number of words), for each recording (see Supplementary Material). The key outcome metrics were the rate of noun production for Story Time Narration, and the ratio of speech duration in each Story Time Free Recall relative to Narration.

#### Object Features

The Object Features semantic task employs an object-feature sentence verification paradigm (e.g. “knife” – “does it have a beak?”). The task manipulated two key variables: semantic complexity (comparing semantically complex living objects with less complex nonliving objects) and distinctiveness (i.e. comparing shared features, which are shared by many concepts and thus more deeply embedded in semantic memory, with distinctive features, which are not shared by many concepts and thus less deeply embedded in semantic memory)^45^. Participants responded to each question by selecting either a “YES” or a “NO” response. The key outcome measure was the mean reaction time to correct responses.

#### Find the Egg

Find the Egg was designed to assess visuospatial working memory^46^, by instructing participants to locate the eggs hiding behind an array of sleeping chickens displayed on the screen. The task consisted of five levels of increasing difficulty, determined by the number of chickens displayed (i.e., 4, 6, 8, 10, or 12). In each level, participants were shown an array of chickens and instructed to tap on one chicken at a time to ‘reveal’ an egg. Once an egg was found, participants dragged it to a basket at the bottom of a screen, concluding that “search” round and starting a new one. Participants were instructed that a chicken could only hide an egg once per level, and thus, tapping a chicken that had already laid an egg was considered an error. Once all eggs in a level were found, the next level started. The task ended if three consecutive errors were committed at any level or if the most difficult level with 12 chickens was successfully completed. The key outcome variable was the highest level reached by the participant.

#### Information Processing Speed

Information Processing Speed was assessed using a modified electronic version of the Symbol Digit Modalities Test (SDMT)^47^ along with an additional digit-digit matching baseline task, to measure visuomotor processing speed and attention. In the baseline task, participants were asked to tap the number on a keyboard that matched the digit displayed in the center of the screen, and it lasted for 15 seconds. In the digit-symbol matching task, participants were presented with a symbol-digit key at the top of the screen, and were instructed to select the corresponding number from a keypad based on the symbol presented in the center of the screen. The digit-symbol task lasted 90 seconds. Each testing session featured novel sequences of correct responses and novel digit-symbol keys. The key outcome metric was the number of correct responses in the digit-symbol task and the mean response time to correct responses in the baseline digit-digit task.

#### 30-Second Dual Task Walking (30sDTW)

This task was designed to measure cognitive-motor-interference by evaluating gait performance during a cognitive dual-task paradigm^48^. Participants placed the study smartphone in a provided running belt and began walking for 30 seconds or approximately 50 meters (54 yards), preferably on even terrain and avoiding U-turns. Simultaneously, they were asked to count backwards by 5 out loud from a randomly pre-specified number. After 30 seconds, the smartphone vibrated to signal the end of the task, at which point participants entered the final number from their countdown on the smartphone screen. Gait was monitored using the smartphone accelerometers and gyroscopes, and counting performance by the smartphone microphones. Step detection was performed using an adaptive step-detection algorithm proposed by Lee et al.^49^, which was previously validated for robust and accurate step-event identification across diverse smartphone orientations. Though the algorithm has not been validated against a gait-lab-based gold standard, in this population, the algorithm has demonstrated the ability to extract reliable gait features^49^. The primary measure for the 30sDTW was median step power (50th percentile). The metric captures the physical effort and walking intensity of each step by calculating the integral of the mean-centered squared acceleration over the step duration. We chose the median value across all walking segments rather than the average to ensure the results were not skewed by a single unusual movement or artifact. In this context, a higher step power reflects more deliberate and purposeful movement by the participants.

#### Speeded Tapping

The Speeded Tapping task assessed fine motor speed and rhythmicity^50^. Participants were instructed to repeatedly tap a large circle on the smartphone screen as quickly as possible using their dominant index finger for 30 seconds, followed by the same task using their non-dominant index finger. The key outcome metric was the mean inter-tap interval.

#### Fairytale Typing Task

The Fairytale Typing Task was designed to assess participants’ psychomotor speed during implicit language processing by asking them to type sentences from the Cinderella fairytale displayed on the screen (see^51^). The task manipulated syntactic complexity, with sentences varying between low and high complexity. Participants were instructed to read each sentence and transcribe it using a keyboard presented at the bottom of the smartphone screen.

Keyboard functionalities such as swipe typing, auto-correction, and landscape mode were disabled to ensure consistency in typing behavior. The key outcome metric was the mean time between keypresses.

#### End-of-Study Questionnaires

A User Experience questionnaire was presented to all participants at the final study visit. This questionnaire asked participants to rate their overall experience and specific aspects of the remote cognitive testing procedure on a 5-point Likert scale (see Supplementary Materials section 1.1.). Study partners completed a survey at the end-of-study visit querying the degree of support they provided participants during the remote testing period (see Supplementary Materials section 1.2.).

### Statistical Analyses

#### Feasibility

Feasibility of smartphone-based remote self-assessment was analyzed descriptively by calculating the proportion of participants who successfully completed the study relative to the total number of participants who started the study. The proportions are presented for each group separately and for the combined group of participants, together with two-sided 95% Clopper-Pearson exact confidence intervals. The following feasibility metrics are additionally reported: the number of participants who withdrew from the study in each group, the average daily time spent performing AD-DAS tasks by group (as collected from the smartphones), and the average level of support required by participants from their study partners.

#### Acceptability

Acceptability of remote cognitive testing was assessed using descriptive summary statistics from the subjective responses from the User Experience Survey. Data from the participants who completed the study were used for this analysis.

#### Adherence

Adherence to smartphone-based remote assessments was defined in two ways: (1) the percentage of remote study days on which participants completed at least one planned task, relative to the expected number of remote days with active testing (i.e., 28 days); and (2) as the percentage of tasks completed by each participant, relative to the total planned number of tasks over the remote monitoring period. Only data from participants who completed the study were included in the analysis, which was pre-specified to be descriptive in nature.

#### Reliability

Test-retest reliabilities were assessed by comparing the median smartphone-based sensor feature values from the first three and the last three valid non-overlapping active task sessions using intraclass correlation coefficients (ICC) with a two-way random effects model with absolute agreement between multiple raters (ICC [2,k])^52^.), moderate (0.50-.75), good (0.75-0.9), and excellent (≥0.9)^53^. Since six data points were required to calculate the ICC (see above), limited data per participant for the Gallery Game short and long delayed memory tasks (i.e., Recognition, Free Recall), and for the Fairy Tale Typing task (due to poor usability of the custom created smartphone keyboard) precluded the calculation of these ICCs.

##### Clinical Validity

###### Association with clinical neuropsychological measures

Data from the median of the last four completed and valid active tasks executed by participants were used for analyses of clinical validity to minimize the potential impact of practice, learning and test sophistication effects. Participants who did not have at least four data points were excluded from this analysis. These analyses used regularized ridge regression models adjusting for age, sex, years of education (six-point ordinal value), and site identifiers (to account for clinical site and language variabilities) as covariates. Regularized ridge regression models were preferred over normal regression to reduce overfitting and to account for multicollinearity between covariates, if any. These models tested for relationships between the AD-DAS outcome metrics and corresponding clinical measures using the ‘glmnet’ package^54^ in R (i.e., [AD-DAS metric] = [_y_ + β_1_[Age] + β_2_[Sex] + β_3_[Education] + β_4_[Site] + β_5_[Clinical Comparator] + [). The resulting adjusted R^2^ for the clinical comparator (interpreted as the square of the semi-partial correlation coefficients between smartphone metric and clinical comparators) and nominal associations (uncorrected p-values) are reported. It is important to note that the classical interpretation of correlation coefficients (i.e., square root of R^2^ values) does not apply, as the present statistic controls for the variance associated with the covariates – unlike standard correlation analyses (e.g., Pearson’s or Spearman’s correlations). Due to the exploratory nature of the study, analyses were descriptive; the nominal p[values are reported without adjustments and should be interpreted as hypothesis-generating rather than confirmatory evidence.

###### Known-group comparisons

Group comparisons tested whether AD-DAS outcome measures differentiated between (a) objectively cognitively impaired (eAD) and each of the not objectively cognitively impaired groups (HC, SCDn, SCDp), and (b) between SCDp and SCDn participants. Proportional odds regression modeled a generalized form of Kruskal-Wallis test with covariate adjustments (i.e., age, sex, education level, site identifier) using the ‘rms’ package^55^ in R. The resulting difference in log odds, along with the sign of the resulting coefficients and nominal associations (uncorrected p-values), were reported. The difference in log-odds is interpreted as the odds of being more or less likely to be in the test group when compared to the reference group, holding all other covariates in the model constant. Thus, the higher the absolute difference in log-odds, the better the separation between the two compared groups. Due to the exploratory nature of the study, the analyses were descriptive; the nominal p[values are reported without adjustments and presented as descriptive markers and not as thresholds for decision making. Group differences should be interpreted using the differences in log odds and their confidence intervals for further hypothesis generation.

Keyboard usability problems with the Fairytale Typing task resulted in data being collected from only 30 of the 123 study participants, only 4 of whom were in the eAD group. Consequently no group comparisons were conducted for this task. All group comparisons were restricted to participants who completed the study.

###### Correlation with whole-brain MRI

Whole-brain voxel-based imaging correlational analyses were conducted on the AD-DAS outcome metrics to explore their relationships with regional brain integrity (atrophy).

Image Preprocessing: Structural (T1) MRI data preprocessing was performed using SPM-12^56^ and the Computational Anatomy Toolbox (CAT)^57^ operated on MATLAB 2021^58^. Image preprocessing consisted of tissue segmentation and spatial registration and were performed based on recommendations from CAT12 manual^57^. Prior to preprocessing, all raw images were visually inspected for artifacts. Each participant’s raw image volume was then segmented into gray matter (GM), white matter (WM) and cerebrospinal fluid using the CAT12 toolbox with standard tissue probability maps (TPMs) provided by SPM. Spatial registration was achieved by applying the high-dimensional DARTEL approach^59^.

<u>Voxel-based morphometry analyses:</u> Whole-brain voxel-based statistical analyses were performed in SPM12 using the spatially normalized and smoothed GM volumes. Second-level general linear models (GLMs) with all participants were specified including age, gender, years of education, and site (which correlates with machine/sequence) as covariates of interest. All non-categorical covariates were mean[centered, while categorical ones were entered as dummy variables. Total intracranial volume (TIV) of each participant was used as a covariate of no interest to adjust for global effects. To explore relationships between smartphone-based cognitive outcome metrics and regional GM integrity (i.e., atrophy), one GLM was built for each key cognitive metric as the covariate of interest. All analyses used standard cluster-level thresholding of p=0.001, and only suprathreshold voxels surviving cluster-level family[wise error (FWE) corrected p-values of 0.05 are reported. These analyses are not reported for the Fairytale Typing Task because of the loss of in particular AD participant data, the Object Features task due to the lack of variability in data, Gallery Game Recognition and Free Recall >24 hours due to the sparsity of data in different recalls, and Story Time Delayed Free Recall due to multiple story exposure during learning. For the VBM analysis of each AD-DAS task, the primary specified outcome measure was used. If this VBM result was negative, then other AD-DAS features were explored; in these cases, the novel metric is explained in the Results section. For comparative purposes, VBMs were also run for the corresponding clinical outcome variables.

## Results

### Feasibility

The overall proportion of participants who completed the study was 97.6%, indicating high feasibility. One healthy volunteer withdrew prematurely due to COVID, and two eAD participants withdrew voluntarily. While the proportions of completers were comparable across groups, more eAD participants withdrew (n=2) compared to all other groups (proportions of completers: HC 97%; SCDn 100%; SCDp 100%; eAD 93%).

### Acceptability

The majority of participants across all subgroups rated the overall experience with remote cognitive monitoring positively, with over 85% expressing acceptance as defined by a score of “good” (46.7%) or “very good” (38.5%). Six (three SCDp and three eAD) participants rated the overall experience as “poor”, and none rated “very poor” (see Table 3 and Supplementary Material section 1.1). Ratings on the acceptability of daily burden, the aggregate burden over the study period, difficulty planning testing sessions, burden of carrying the study smartphone throughout the day, and ease of understanding the instructions all had a mean positive rating. The mean rating for the difficulty of using the running belt for the gait task was “not difficult” for the HC group (response option 1), and moderately difficult for the remaining groups (response option 2).

**Table 3.**
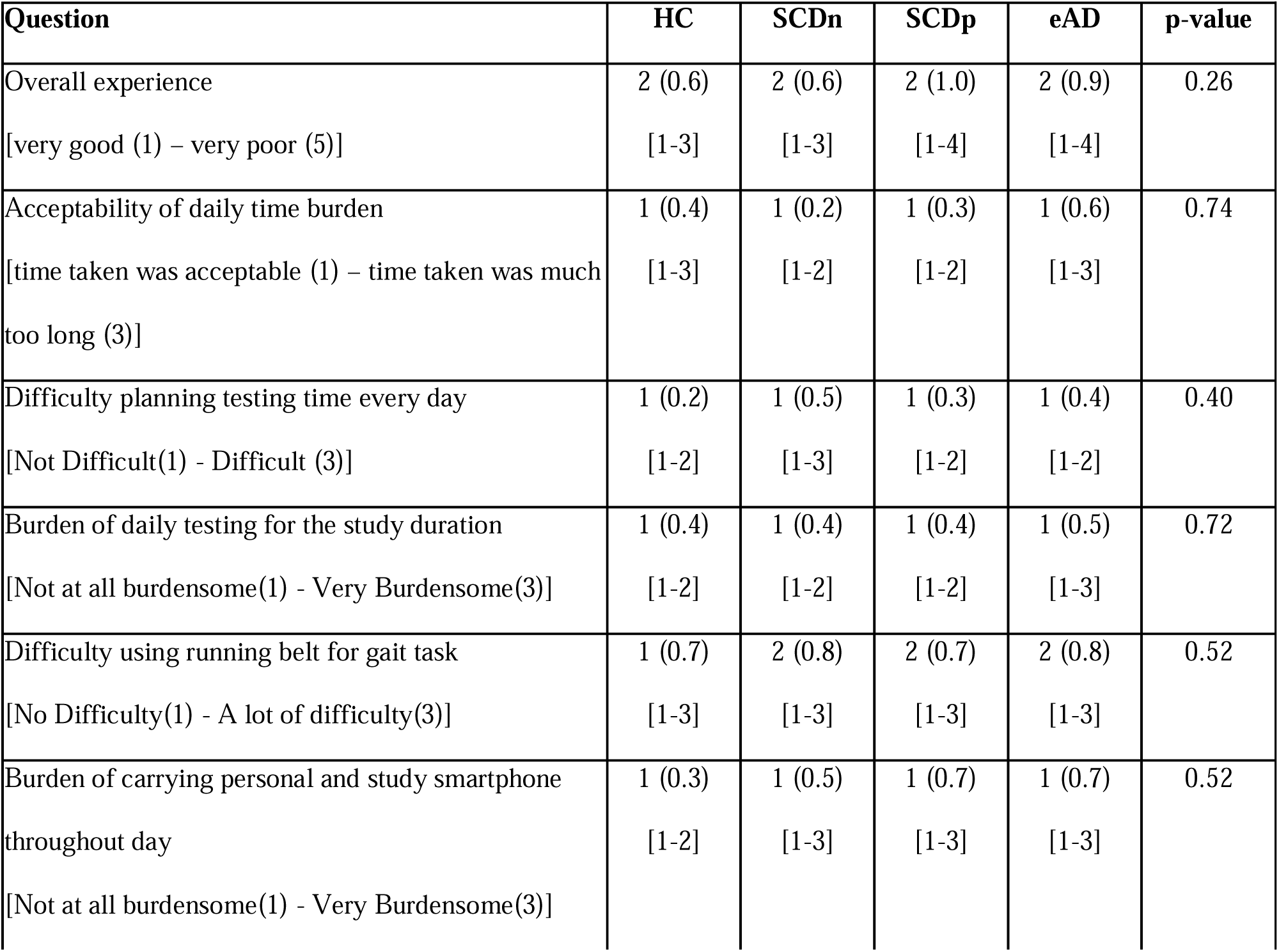
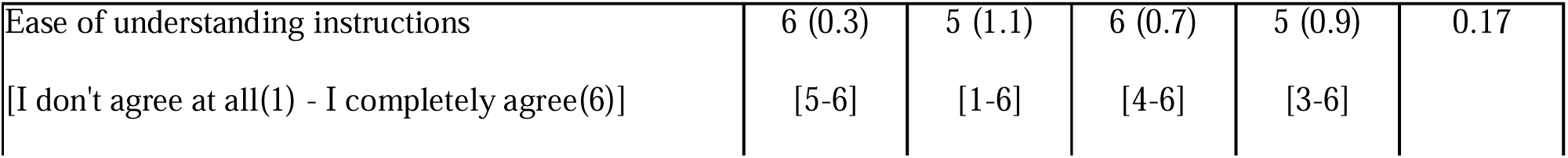
Summary of participant experience with AD-DAS cognitive testing (Mean (Standard deviation) [range]) based on responses to a questionnaire. Comparison of distributions across groups was performed using Fisher exact test.

The overall median time spent performing the remote assessments was 14.5 minutes per day (see Figure 1a), with the HC participants requiring the least (13.2 minutes) and the eAD participants the most (16.6 minutes) amount of time. According to study partners, the proportion of participants who required assistance in completing the AD-DAS tasks (“Could the study participant do the tasks without assistance? (Yes/No)”) were as follows: HC 0/32; SCDn 1/31; SCDp 4/30; SCDp and eAD 9/30.

**Figure 1.**
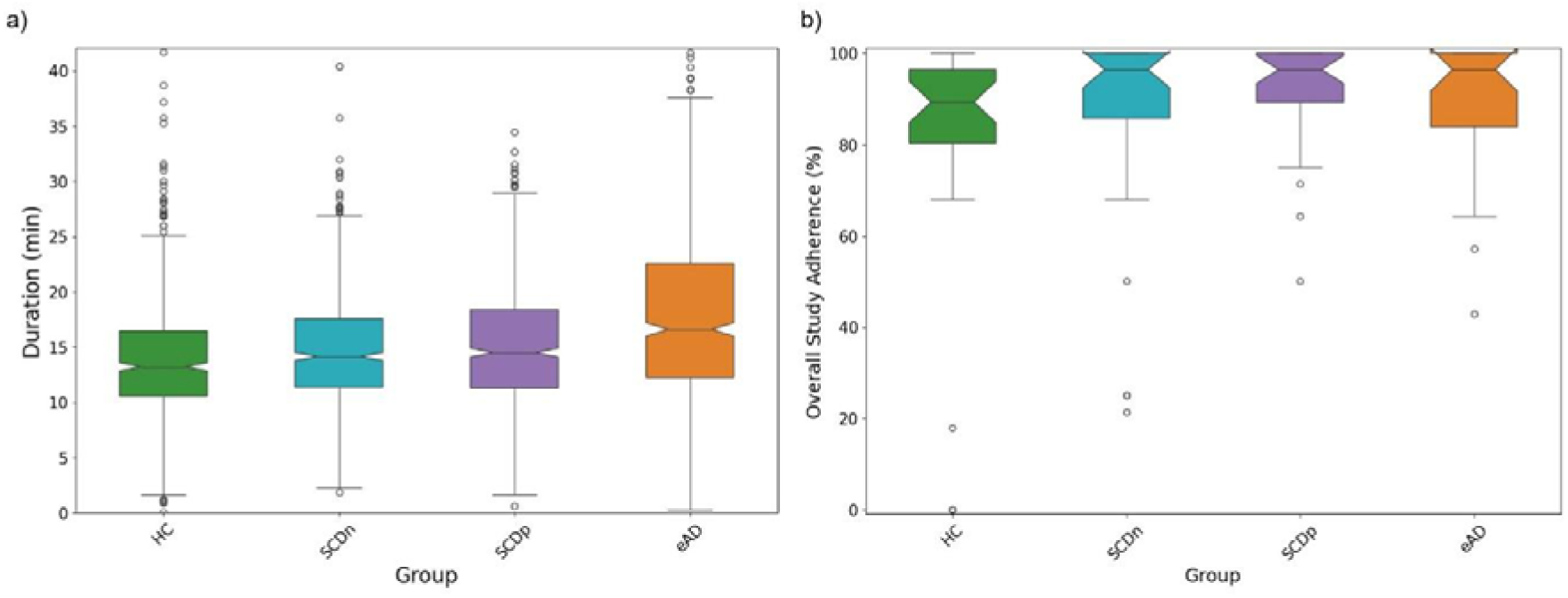
Daily AD-DAS burden and overall adherence. (a) Time (in minutes) per day required to complete the AD-DAS battery. (b) Overall study adherence (in %) to AD-DAS remote assessments.

### Adherence

The median adherence to the remote testing protocol was circa 96% among participants who successfully completed the study, i.e., around 27–28/28 days (see Figure 1b). However, a lower proportion of HC participants completed the tests (89.3%) compared to the remaining subgroups. The average adherence to individual Active Tasks was high (96.4%). Individual Active Task adherence ranged from 77% (Gallery Game Recall) to 100% (Tilt Task) (see Supplementary Table 1).

### Reliability

AD-DAS features demonstrated moderate to excellent^53^ test-retest reliabilities (ICCs; see Table 4). Mean ICCs ranged from 0.53 (speeded tapping with the non-dominant hand) to 0.91 (speeded tapping with the dominant hand).

**Table 4.**
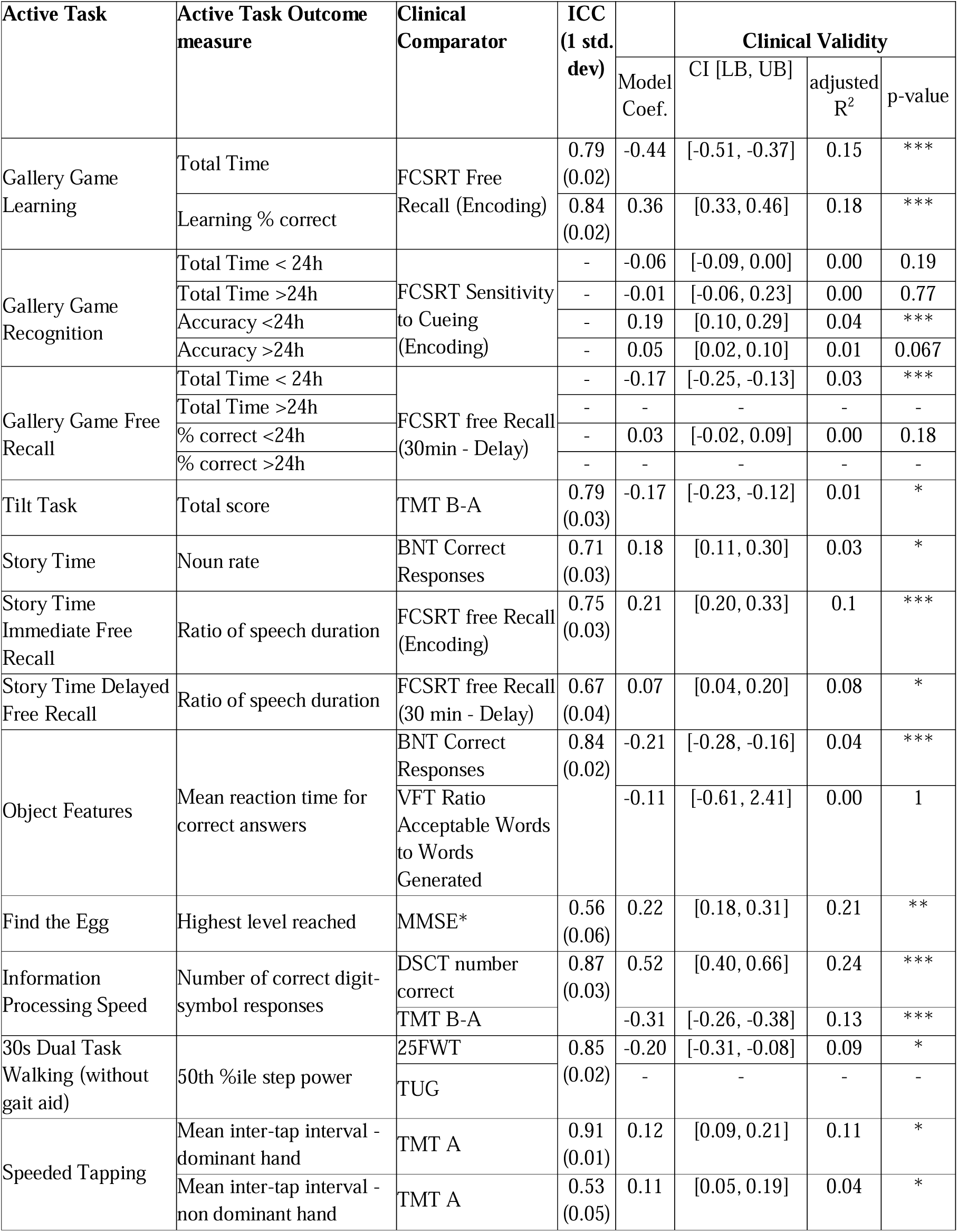
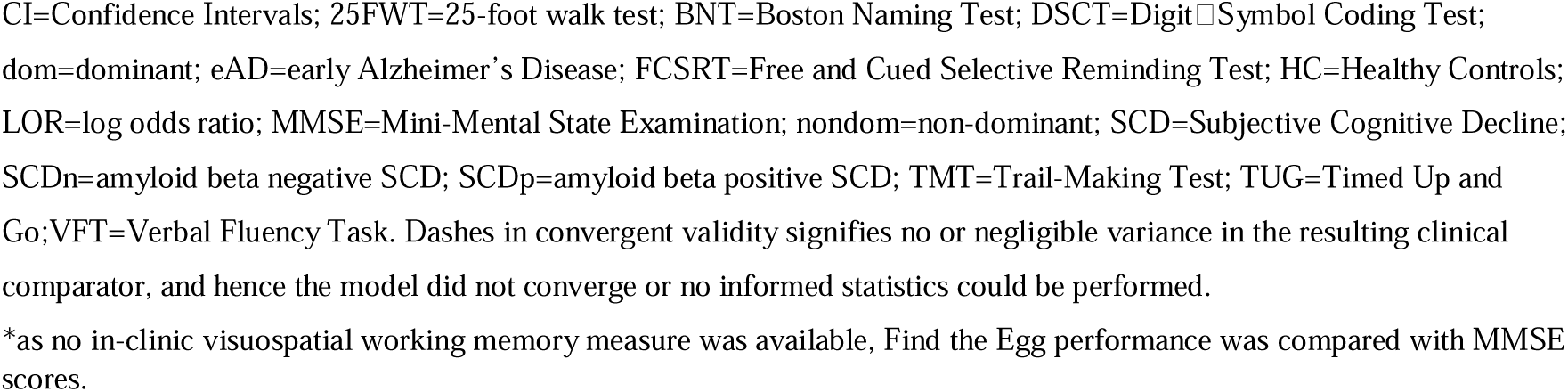
Reliability and preliminary clinical validity (comparisons with clinical comparators) of the AD-DAS active test outcome measures (* unadjusted p-value ≤ 5×10-2; ** unadjusted p-value ≤ 5×10-3; and *** unadjusted p-value ≤ 5×10-4).

### Clinical Validity

#### Association with clinical neuropsychological measures

The model coefficients, adjusted R^2^ (interpreted as the square of the semi-partial correlation coefficients), and unadjusted p-values indicate the direction, strength, and nominal association of the convergent validity findings, respectively (cf. Table 4 and Supplementary Figure 1). At least one metric from all AD-DAS tasks was associated with the clinical comparator in the expected direction, with the exception of the Fairytale task (results not shown due to data missingness). For example, total time during Gallery Game Learning was negatively associated with FCSRT free recall during encoding (R^2^ = 0.15, p < 4.1e-5), while the percentage of correctly learned objects positively associated with FCSRT free recall (R^2^ = 0.18, p < 1.9e-4). Gallery Game Recognition (both <24H and >24h) accuracy was marginally positively associated with FCSRT sensitivity to cueing during encoding (R^2^ = 0.04 for <24h, R^2^ = 0.01 for >24h). Tilt Task total scores negatively associated with TMT B-A, suggesting that participants with more efficient task switching capabilities advanced further in the Tilt Task. The noun rate in Story Time Narration positively associated with BNT scores, while speech duration ratio in Story Time Delayed Free Recall associated with FCSRT free recall collected after 30 min delay. However, the speech duration ratio in Story Time Immediate Free Recall showed no association with FCSRT free recall (encoding). All the other assessments, including Find the Egg, Information Processing Speed, 30-sec Dual Task Walking, and Speeded Tapping showed nominal associations with their respective comparators in the expected directions. Overall, the effect sizes of these comparisons ranged from low (adjusted R^2^ = 0.01) to moderate (adjusted R^2^ = 0.24), supporting preliminary validity.

#### Known-group comparisons

Group comparisons were conducted across HC, SCDn, SCDp and eAD groups to evaluate the preliminary clinical validity of AD-DAS metrics (Figure 2, Table 4 and Supplementary Figure 2). The majority of AD-DAS metrics differentiated the eAD from other groups, with the exception of 30sDTW gait speed and non-dominant hand mean tapping interval in the Speeded Tapping task, which differentiated eAD from HC and SCDp but not from SCDn.

**Figure 2.**
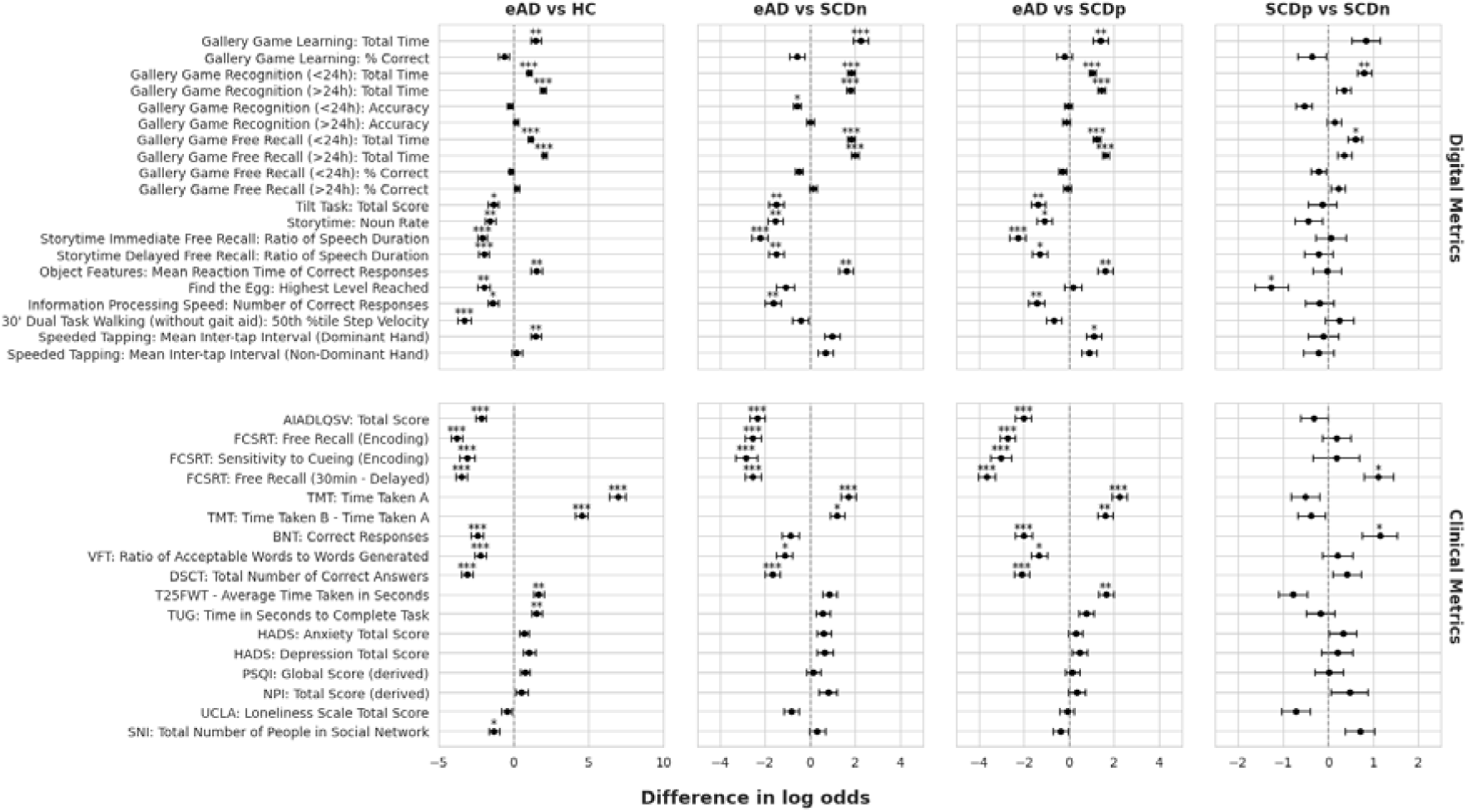
Clinical validity of AD-DAS metrics (top row) and the in-clinic standard neuropsychological assessments (bottom row) to differentiate between known groups (columns). Each column represents a unique pairwise comparison. The x-axis indicates the mean differences in the log of odds between two groups, and the y-axis represents the various assessments and their key metrics of interest. Descriptive statistics for these comparisons are denoted with unadjusted p-values for transparency and not as thresholds for decision-making (* ≤ 5×10-2; ** p-value ≤ 5×10-3; and *** ≤ 5×10-4.)

Among the clinical scales, AIADLQSV, FCSRT, TMT, BNT, VFT and DSCT differentiated groups. T25FWT and TUG differentiated eAD from HC but not other groups. These results are summarized in Figure 2 and Supplementary Table 2.

SCDp individuals, i.e. those at higher risk to progress to MCI, demonstrated greater impairments in Gallery Game Recognition (<24h), Gallery Game Free Recall (<24h), and Find the Egg highest level reached compared to SCDn participants (see Figure 2). Among the classical neuropsychological scales (which were also not used for SCD participant inclusion/exclusion), FCSRT free recall after 30 min delay and BNT showed nominally SDCn-SCDp differences (see Figure 2). These findings provide preliminary evidence for the sensitivity of AD-DAS metrics to detect subtle cognitive changes in the early spectrum of AD.

##### VBM whole-brain MRI associations

The results of the VBM analyses of digital variables are shown in Figure 3 and both digital and clinical VBM results in Supplementary Table 3. The more time taken to reach criterion in the Gallery Game Learning task was associated with reduced gray matter (GM) volumes in the right middle occipital lobe (extending into the inferior occipital, lingual gyrus and posterior inferior temporal lobe), and bilateral perirhinal cortices (encompassing the temporal poles, entorhinal cortices, fusiform gyri and inferior temporal lobes). Similar results were found with FCSRT free recall (encoding), with associations in bilateral hippocampi and anterior parahippocampal and inferior and middle temporal cortices. These regions are implicated in the visual processing and episodic learning^60^. While no clusters were identified for Gallery Game Recognition task accuracy or completion time, the hit rate (>24h) was associated with gray matter volumes in the left thalamus. The lack of association with perirhinal cortex and hippocampal integrity was surprising; however, the association with thalamus is consistent with its hypothesized role as an object recognition output structure^61^. A related cognitive measure, FCSRT sensitivity to cueing, was associated with bilateral orbitofrontal, and inferior and middle temporal regions, in line with previous reports^62^. Longer completion times for the Gallery Game Free Recall (<24 hours) was associated with reduced GM volumes in the left perirhinal and entorhinal cortices, extending into the middle and superior temporal poles, supporting object processing and memory recall^60^, as expected. Similar findings were evident in the FCSRT delayed free recall (Supplementary Table 3).

**Figure 3.**
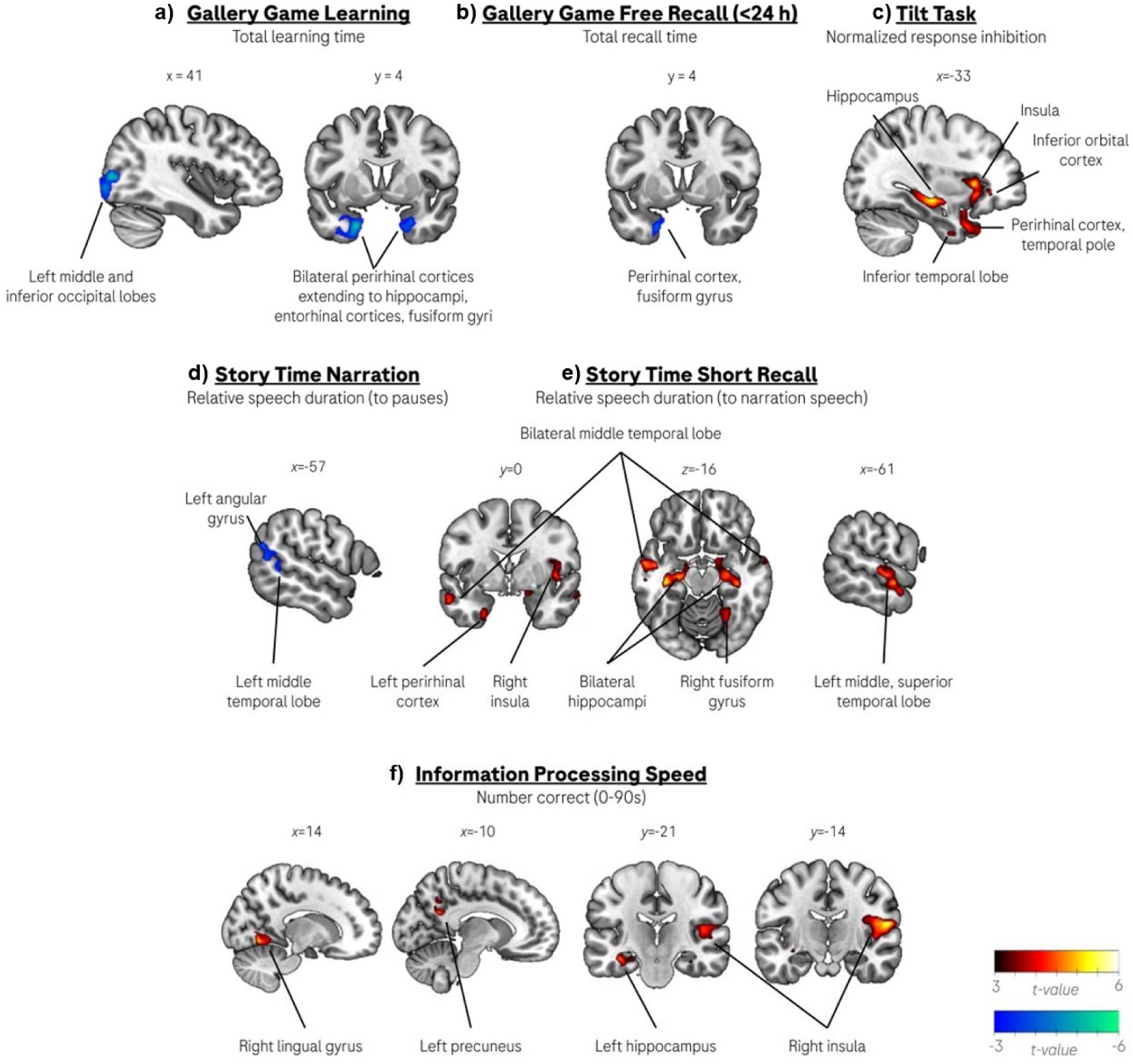
Regions of the brain where select AD-DAS task performance correlated with estimates of gray matter integrity (see Supplementary Table 3 for full details).

While neither total scores on the Tilt Task nor TMT B-A showed associations with GM, accuracy on Tilt Task response inhibition trials (normalized by non-inhibition trials) was associated with clusters in regions associated with visual processing (calcarine sulcus), complex object processing (left anterior inferior temporal lobe), memory (bilateral hippocampi), and, notably, response inhibition (left inferior orbital cortex, bilateral insulae). Thus, these results are in line with core response inhibition structures working together with Tilt Task specific visual working memory networks, as in other tasks requiring the inhibition of pre-potent responses^63^.

The Story Time Narration rate of noun production, BNT and VFT total numbers of correct answers, all did not yield supra-threshold clusters. However, the ratio of speech to pause durations during Story Time Narration was associated with reduced GM volumes in the left cerebellum (extending into the left fusiform gyrus) and the left angular gyrus (extending into the supramarginal gyrus and middle temporal gyrus), supporting language production and semantic processing^64^. Moreover, lesion and functional imaging studies of the cerebellum have demonstrated that this structure supports numerous language functions including production^65^. The ratio of speech duration in Story Time Immediate Free Recall to that in Story Time Narration task was associated with GM integrity of a complex set of brain regions in language processing associate areas (left middle, superior temporal lobes and Rolandic operculum)^64^, the insula (coordinate language processing and production)^66^ and memory related regions (anteromedial temporal lobe including the bilateral hippocampus and perirhinal and entorhinal cortices).

Higher levels reached on the Find the Egg task were associated with GM volumes in the left cerebellum, consistent with its role in visuospatial working memory^67^. We note that no clinical measures of visuospatial working memory were available for VBM comparison.

Reduced performance on the Information Processing Speed task (i.e., lower number of correct digit-symbol responses in 90 seconds) was associated with GM atrophy in the left hippocampus and precuneus extending into the cuneus, as well as the right insula/Rolandic and temporal operculum (STG)/Heschl’s gyrus and lingual gyrus. A recent meta-analysis of functional imaging studies of Symbol Digit Modalities Test (SDMT) demonstrated that the lingual gyrus and precuneus/cuneus were commonly found to be activated during SDMT performance, consistent with the former structure’s role in visual pattern recognition and visual attention, and the latter’s association with visuospatial imagery, spatial attention and memory retrieval^68^. There were no suprathreshold clusters in VBM with the DSCT total number correct.

The observed task-specific correlations between GM atrophy and digital task performance metrics recapitulate structure-function relationships established in tasks assessing similar domains of cognition, and provide a biomarker-based validation of the relevance of these digital measures to neurological and cognitive integrity in the early AD spectrum.

## Discussion

This study provides preliminary evidence that daily, remote, smartphone-based assessments of a broad array of cognitive functions with the AD-DAS over a 28-day testing period is feasible (97.6%) and acceptable to HC, SCD and eAD participants, with an average adherence of 96% to the remote testing protocol. The majority of participants across all subgroups rated the experience with remote cognitive monitoring positively, with an average rating of >85% good/very good. AD-DAS metrics demonstrated moderate to excellent^53^ test-retest reliability (0.53–0.91), and showed modest associations with their in-clinic neuropsychological comparators (adjusted R^2^ from 0.01 to 0.24), and distinguished between cognitively impaired and cognitively non-impaired groups (i.e., eAD vs. HC, SCDn, SCDp), with confidence intervals indicating preliminary validation. The relatively lower magnitude of the adjusted R^2^values reflects both covariate adjustments and limited sample sizes. Additionally, SCDp participants performed worse than SCDn participants on three AD-DAS tasks (see also^69^).

VBM findings were broadly consistent with the expected structure-function relationships, demonstrating relationships between AD-DAS outcome measures and volumes in the expected brain regions, and the lack thereof in unexpected brain regions, providing further convergent validity of the AD-DAS tasks. The present results are notable in demonstrating robust psychometric properties and acceptance and adherence, with the relatively broad scope of remote cognitive testing resulting in longer daily testing sessions (ca. 11 minutes) and longer span of testing days (26/28 days).

AD-DAS metrics were associated with the expected neuropsychological test measures obtained in the clinic. We note that case-control differences in Gallery Game performance were not as pronounced as expected. Gallery Game differed from standard, fixed-trial episodic learning paradigms by employing an incremented[difficulty approach^70^ whereby at first two, and up to six objects were pseudorandomly and repeatedly presented as many times as necessary to correctly recall each stimulus association (i.e., direction-object association) on at least five of the last six presentations^42^. We speculate that this design may have enabled participants – including eAD patients – to benefit from a ‘spacing effect’ hypothesized to rely on the basal ganglia habit learning circuit^71^. The spacing effect describes the phenomenon that the memorability of items which are repeatedly presented for learning is increased when the learning items are presented in a spaced fashion (i.e., typically spaced in time but also shown when spaced with intervening items), compared to when the items are consecutively presented for learning in bulk^72^.

AD-DAS cognitive outcome measures differentiated eAD from HC, and in the majority of cases also eAD from SCDn and SCDp groups, as expected. Moreover, the SCDp group performed worse than the SCDn group on three AD-DAS tasks: Gallery Game Recognition and Gallery Game Free Recall (both <24h), and visuospatial working memory as measured by the Find the Egg task. These findings are in line with those from Vogel et al.^69^, who demonstrated that SCD severity and amyloid PET positivity independently predicted subsequent cognitive decline in episodic memory and global cognitive functioning, while more severe SCD predicted faster decline in working memory, independent of amyloid PET status. This distinction indicates that while the SCDn-SCDp difference in episodic memory may reflect a heightened risk to progress to MCI/AD, the predictive significance of working memory impairments require further investigation (see also^73^).

Similarly robust feasibility, reliability and clinical validity have been reported for other remote cognitive testing solutions, notably the CBB^19^ and ARC^22^. Remote CBB assessments each lasting about 20 minutes were highly feasible (98% completion rate) and showed good usability^18^. Three-month test-retest reliability (ICCs) ranged from 0.76 to 0.93^74^. Clinical validity was supported by correlations with ADAS-Cog and MOCA (r ≥ 0.4), and six of the 16 baseline measures differentiated healthy controls from MCI (Cohen’s d ≥ 0.3)^19^.

ARC comprises brief tasks administered multiple times per day (i.e., 3-5) for several minutes each, and has shown feasibility, acceptability over 7 days, and reliabilities above 0.85 over 6-12 months^22^. ICCs for each ARC task increased as a function of the number of testing sessions aggregated, reaching ≥ 0.90 with 5 symbol search and n-back testing sessions, and with 15 dot memory sessions^21^. ARC measures also correlates with classical cognitive measures (i.e., r = 0.22 - 0.57), and the ARC composite with CSF, PET and MRI biomarkers (i.e., r = 0.11 - 0.29)^22^.

Thus, several feasible, acceptable and partially validated remote cognitive testing batteries for early AD are available, each with distinct advantages regarding brevity, testing frequency, breadth of cognitive domains and inclusion of long-term episodic memory. Single daily testing sessions of the CBB (∼ 17 minutes^19^) is comparable to that of the AD-DAS, whereas ARC sessions are shorter (∼ 3 minutes) but administered 3-5 times per day. Reliability estimates from aggregated ARC^21^ sessions and repeated CBB administrations are comparable, and broadly consistent with those observed for AD-DAS. All the three batteries show preliminary associations with in-clinic cognitive measures and differentiate groups across the AD spectrum. They differ in the range of cognitive tests administered (three in CBB and ARC, nine in AD-DAS towards supporting differential diagnostic use cases), cognitive domains tested, and in the inclusion of a long-term episodic memory task (AD-DAS).

Accordingly, selection of an appropriate DHT should be guided by the specific use case and study design constraints.

Whole brain, voxel-based morphometry analyses were conducted to determine whether performance on select smartphone-based cognitive assessments correlated with the integrity of brain regions hypothesized to underpin the associated functions. Importantly, whole-brain (as opposed to region of interest) approaches were used to confirm the specificity of the relationships, i.e. that task performance correlated with atrophy in the expected, but not in the unexpected, brain regions. Thus, VBM analyses offered an orthogonal source of content validation for the AD-DAS cognitive outcome metrics. These results largely confirmed the clinical validity findings, as smartphone-based cognitive task performance correlated with atrophy of the convergent (but not divergent) brain structures. In addition, the patterns of VBM results were comparable across digital and clinical measures.

Considerations of brain-behavior relationships and AD neuropathology can inform the design and selection of cognitive tools for the detection of individuals in the earliest stages of the disease, as was the case for the AD-DAS battery. The pathology most closely associated with cognitive disturbances in AD, i.e., intracellular neurofibrillary pathology^75^, begins cortically in the medial perirhinal cortex, before spreading medially to the entorhinal cortex and hippocampus, and thereafter laterally to the anterior fusiform gyri and posteriorly in the parahippocampal cortex and beyond^76^. The pattern of cortical thinning in different stages of the AD spectrum mirrors this progression^77^, as does the temporal emergence of cognitive changes in prodromal AD. For example, the longitudinal BAsel Study on the ELderly (BASEL study) demonstrated that the verbal episodic delayed recall performance was the first measured cognitive function to decline during the prodromal course of AD, eight years preceding diagnoses of MCI due to AD, as confirmed on average 4 years later^12^. Notably, MMSE scores from the MCI due to AD group only differed from the control cohort at the timepoint of the MCI diagnosis^12^. More recent findings demonstrating perirhinal involvement in the recognition and recall of visually and semantically complex objects may imply that tasks which strongly invoke those cognitive demands could offer a further opportunity to enhance the detection of prodromal AD^78,79^.

Limitations of the present study include the modest sample size, cross-sectional design, and limited demographic diversity; thus, the present results require validation in an independent and larger study to generate diagnostic accuracy data for individuals on the preclinical and early AD spectrum. Such a study would further inform test selection for this case-finding use case, towards minimizing the time required to complete the tasks. An additional limitation is the lack of detailed participant debriefing on individual cognitive tasks, which would have informed usability improvements.

Future directions, as for any novel cognitive tool, would include the collection of large normative datasets. These are critical to appropriately control for the influence of demographic variables on cognitive performance and increase the precision of case-finding. Ideally, these normative studies would be longitudinal in nature to enable norms for change critical to diagnostic requirements to demonstrate a change from a previous level of functioning^80^. For use in clinical drug development, longitudinal studies quantifying relationships between changes in the DHT with changes in primary (e.g. CDR-SOB^81^) or key secondary (e.g. PACC5^82^) outcome measures, and the ability of short-term DHT changes to predict long-term changes in the clinical measures, would be important. Future research should also consider longitudinally studying healthy agers and individuals with prodromal and early AD with several different remote testing solutions. Such a study would provide the data required to identify the optimal combination of outcome measures *across* solutions to differentiate groups, predict progression of key clinical outcome measures, and track cognitive decline over time.

In the context of disease-modifying therapies for AD, the ultimate aim is to intervene in a stage of the disease when the least amount of neurodegeneration has taken place, i.e., to maximize the likelihood of neuronal and functional sparing with positive impacts on quality of life. Early intervention is also expected to have positive socioeconomic impacts. Indeed, a Markov model simulation by Budd and colleagues^13^ demonstrated that earlier detection and treatment of individuals with disease-modifying compounds increased time spent in mild predementia stages (3.2 to 4.2 years), decreased time in moderate to severe AD stages (2.6 to 2.2 years), increased time spent in the community (4.4 to 5.4 years) and decreased time in long-term care (1.3 to 0.9 years). Notably, a significant impact of screening accuracy on socioeconomic savings was reported. The remote measurement of cognitive functioning may represent a feasible and efficient alternative to identifying individuals at early timepoints on the AD continuum. The AD-DAS was designed to address some of the key anticipated challenges in enabling access to disease-modifying therapies: assessment of key cognitive domains known to be affected in preclinical eAD, in particular long delay episodic memory^12^, the potential for multiple testing sessions to increase the reliability of the outcome measures, the testing of a broad range of cognitive and motor functions to contribute to differential diagnostic considerations, and in theory a very broad reach (i.e., anyone with a smartphone) including those not living near specialist centers^2^. This contrasts with the more focused ambulatory assessments like ARC^22^ and M2C2^4,21^ platforms. The present findings indicate that the AD-DAS may represent a candidate tool fulfilling these aims, pending confirmatory validation.

## Data Availability

For eligible studies qualified researchers may request access to individual participant-level clinical data through a data request platform. At the time of writing this request platform is Vivli (https://vivli.org/ourmember/roche/). For up-to-date details on Roche′s Global Policy on the Sharing of Clinical Information and how to request access to related clinical study documents, see here: https://go.roche.com/data_sharing. Anonymized records for individual participants across more than one data source external to Roche cannot, and should not, be linked due to a potential increase in risk of patient re–identification.

https://go.roche.com/data_sharing

## Acknowledgements

This work was funded by F. Hoffmann-La Roche Ltd. We extend our appreciation to the participants, their study partners, and site staff. We also acknowledge the invaluable insights and guidance provided by Dr. Irene Meier, Dr. Kaycee Sink, Dr. Gregory Klein, Dr. Tobias Bittner, Dr. Luka Kulic, Dr. Geoffery Kerchner, Ms. Kami Kosek and the Roche expert reviewers.

## Author contributions

KIT and TMP wrote the first draft; KIT, FO, MGVC, SR-R, DW, MB and TMP designed the study; KIT, AMW, ITK, MV, GK, NH, BS, MGVC, WA, SR-R, EP, DW, MB and TMP aided in the conduct of the study; AMW, ITK, GK, NH, EAA, SH, RU and TMP analyzed the data; KIT, AMW, ITK, NH, EAA, CHC, SH, RU, DW, MB and TMP interpreted the results; KIT, AMW, ITK, NH, EAA, CHC, SH, RU, DW, MB and TMP revised the manuscript. KIT confirms that all authors have read and approved the manuscript.

## Data availability

For eligible studies qualified researchers may request access to individual participant-level clinical data through a data request platform. At the time of writing this request platform is Vivli (https://vivli.org/ourmember/roche/). For up-to-date details on Roche’s Global Policy on the Sharing of Clinical Information and how to request access to related clinical study documents, see here: https://go.roche.com/data_sharing. Anonymized records for individual participants across more than one data source external to Roche cannot, and should not, be linked due to a potential increase in risk of patient re-identification.

## References

1. Brookmeyer, R., Johnson, E., Ziegler-Graham, K. & Arrighi, H. M. Forecasting the global burden of Alzheimer’s disease. Alzheimers Dement. 3, 186–191 (2007).

2. Belder, C. R. S., Schott, J. M. & Fox, N. C. Preparing for disease-modifying therapies in Alzheimer’s disease. Lancet Neurol. 22, 782–783 (2023).

3. Morley, J. E. et al. Brain health: the importance of recognizing cognitive impairment: an IAGG consensus conference. J. Am. Med. Dir. Assoc. 16, 731–739 (2015).

4. Singh, S. et al. Ecological Momentary Assessment of Cognition in Clinical and Community Samples: Reliability and Validity Study. J. Med. Internet Res. 25, e45028 (2023).

5. Ashford, J. W. et al. Now is the Time to Improve Cognitive Screening and Assessment for Clinical and Research Advancement. J. Alzheimers Dis. JAD 87, 305–315 (2022).

6. Öhman, F. et al. Unsupervised mobile app-based cognitive testing in a population-based study of older adults born 1944. Front. Digit. Health 4, 933265 (2022).

7. Mitchell, A. J. & Malladi, S. Screening and case finding tools for the detection of dementia. Part I: evidence-based meta-analysis of multidomain tests. Am. J. Geriatr. Psychiatry Off. J. Am. Assoc. Geriatr. Psychiatry 18, 759–782 (2010).

8. Brown, J. The use and misuse of short cognitive tests in the diagnosis of dementia. J. Neurol. Neurosurg. Psychiatry 86, 680–685 (2015).

9. Cannon, P. & Larner, A. J. Errors in the scoring and reporting of cognitive screening instruments administered in primary care. Neurodegener. Dis. Manag. 6, 271–276 (2016).

10. Miller, D. S., Wang, Xi. & Kott, A. Exploring the causes of MMSE scoring and administration errors in the clinical trial screening period: Neuropsychiatry and behavioral neurology/treatment development and clinical trials. Alzheimers Dement. 16, e045353 (2020).

11. Beishon, L. C. et al. Addenbrooke’s Cognitive Examination III (ACE-III) and mini-ACE for the detection of dementia and mild cognitive impairment. Cochrane Database Syst. Rev. 2019, (2019).

12. Mistridis, P., Krumm, S., Monsch, A. U., Berres, M. & Taylor, K. I. The 12 Years Preceding Mild Cognitive Impairment Due to Alzheimer’s Disease: The Temporal Emergence of Cognitive Decline. J. Alzheimers Dis. JAD 48, 1095–1107 (2015).

13. Budd, D., Burns, L. C., Guo, Z., L’italien, G. & Lapuerta, P. Impact of early intervention and disease modification in patients with predementia Alzheimer’s disease: a Markov model simulation. Clin. Outcomes Res. CEOR 3, 189–195 (2011).

14. Öhman, F., Hassenstab, J., Berron, D., Schöll, M. & Papp, K. V. Current advances in digital cognitive assessment for preclinical Alzheimer’s disease. Alzheimers Dement. Diagn. Assess. Dis. Monit. 13, (2021).

15. Román, G. C. & Royall, D. R. Executive control function: a rational basis for the diagnosis of vascular dementia. Alzheimer Dis. Assoc. Disord. 13 Suppl 3, S69–80 (1999).

16. Zuidersma, M. et al. Temporal dynamics of depression, cognitive performance and sleep in older persons with depressive symptoms and cognitive impairments: a series of eight single-subject studies. Int. Psychogeriatr. 34, 47–59 (2022).

17. Stricker, N. H. et al. A novel computer adaptive word list memory test optimized for remote assessment: Psychometric properties and associations with neurodegenerative biomarkers in older women without dementia. Alzheimers Dement. Amst. Neth. 14, e12299 (2022).

18. Perin, S. et al. Unsupervised assessment of cognition in the Healthy Brain Project: Implications for web-based registries of individuals at risk for Alzheimer’s disease. Alzheimers Dement. N. Y. N 6, e12043 (2020).

19. Edgar, C. J. et al. Pilot Evaluation of the Unsupervised, At-Home Cogstate Brief Battery in ADNI-2. J. Alzheimers Dis. JAD 83, 915–925 (2021).

20. Stawski, R. S., MacDonald, S. W. S. & Sliwinski, M. J. Measurement Burst Design. in The Encyclopedia of Adulthood and Aging (ed. Whitbourne, S. K.) 1–5 (Wiley, 2015). doi:10.1002/9781118521373.wbeaa313.

21. Sliwinski, M. J. et al. Reliability and Validity of Ambulatory Cognitive Assessments. Assessment 25, 14–30 (2018).

22. Nicosia, J. et al. Unsupervised high-frequency smartphone-based cognitive assessments are reliable, valid, and feasible in older adults at risk for Alzheimer’s disease. J. Int. Neuropsychol. Soc. 29, 459–471 (2023).

23. Jessen, F. et al. A conceptual framework for research on subjective cognitive decline in preclinical Alzheimer’s disease. Alzheimers Dement. J. Alzheimers Assoc. 10, 844–852 (2014).

24. Pike, K. E., Cavuoto, M. G., Li, L., Wright, B. J. & Kinsella, G. J. Subjective Cognitive Decline: Level of Risk for Future Dementia and Mild Cognitive Impairment, a Meta-Analysis of Longitudinal Studies. Neuropsychol. Rev. 32, 703–735 (2022).

25. Jessen, F. et al. The characterisation of subjective cognitive decline. Lancet Neurol. 19, 271–278 (2020).

26. An, R. et al. Predictors of progression from subjective cognitive decline to objective cognitive impairment: A systematic review and meta-analysis of longitudinal studies. Int. J. Nurs. Stud. 149, 104629 (2024).

27. Grober, E. & Buschke, H. Genuine memory deficits in dementia. Dev. Neuropsychol. 3, 13–36 (1987).

28. Army Individual Test Battery. Trail Making Test. (1984).

29. Monsch, A. U. et al. Comparisons of verbal fluency tasks in the detection of dementia of the Alzheimer type. Arch. Neurol. 49, 1253–8 (1992).

30. Wechsler, D. Wechsler Adult Intelligence Scale--Third Edition. 10.1037/t49755-000 (2019).

31. Kaplan, E. F., Goodglass, H. & Weintraub, S. The Boston Naming Test. (Veterans Administration Medical Center, Boston, 1978).

32. Rattanabannakit, C. et al. The Cognitive Change Index as a Measure of Self and Informant Perception of Cognitive Decline: Relation to Neuropsychological Tests. J. Alzheimers Dis. JAD 51, 1145–1155 (2016).

33. Risacher, S. L. et al. APOE effect on Alzheimer’s disease biomarkers in older adults with significant memory concern. Alzheimers Dement. J. Alzheimers Assoc. 11, 1417–1429 (2015).

34. Folstein, M. F., Folstein, S. E. & McHugh, P. R. ‘Mini Mental State’ - a practical method for grading the cognitive state of patients for the clinician. J. Psychiatry Res. 12, 189–98 (1975).

35. Bravo, G. & Hébert, R. Age- and education-specific reference values for the Mini-Mental and modified Mini-Mental State Examinations derived from a non-demented elderly population. Int.

36. J. Geriatr. Psychiatry 12, 1008–1018 (1997).

36. Morris, J. C. The Clinical Dementia Rating (CDR): current version and scoring rules. Neurology 43, 2412–2414 (1993).

37. Jack, C. R. et al. NIA-AA Research Framework: Toward a biological definition of Alzheimer’s disease. Alzheimers Dement. J. Alzheimers Assoc. 14, 535–562 (2018).

38. Thomas-Anterion, C., Honore-Masson, S. & Laurent, B. The cognitive complaint interview (CCI). Psychogeriatrics 6, S18–S22 (2006).

39. Jutten, R. J. et al. Detecting functional decline from normal aging to dementia: Development and validation of a short version of the Amsterdam IADL Questionnaire. Alzheimers Dement. Diagn. Assess. Dis. Monit. 8, 26–35 (2017).

40. Hauser, S. L. et al. Intensive immunosuppression in progressive multiple sclerosis. A randomized, three-arm study of high-dose intravenous cyclophosphamide, plasma exchange, and ACTH. N. Engl. J. Med. 308, 173–180 (1983).

41. Kear, B. M., Guck, T. P. & McGaha, A. L. Timed Up and Go (TUG) Test: Normative Reference Values for Ages 20 to 59 Years and Relationships With Physical and Mental Health Risk Factors. J. Prim. Care Community Health 8, 9–13 (2017).

42. Lancaster, C. et al. Gallery Game: Smartphone-based assessment of long-term memory in adults at risk of Alzheimer’s disease. J. Clin. Exp. Neuropsychol. 1–15 (2020) doi:10.1080/13803395.2020.1714551.

43. Stroop, J. R. Studies of interference in serial verbal reactions. J. Exp. Psychol. 18, 643–662 (1935).

44. Wechsler, D. A Standardized Memory Scale for Clinical Use. J. Psychol. 19, 87–95 (1945).

45. Taylor, K. I., Moss, H. E. & Tyler, L. K. The Conceptual Structure Account: a cognitive model of semantic memory and its neural instantiation. in The Neural Basis of Semantic Memory (eds Hart, J. & Kraut, M.) 265–301 (Cambridge University Press, Cambridge, UK, 2007).

46. Lezak, M., Howieson, D. B., Bigler, E. D. & Tranel, D. Neuropsychological Assessment. (Oxford University Press, 2012).

47. Silva, P. H. R., Spedo, C. T., Barreira, A. A. & Leoni, R. F. Symbol Digit Modalities Test adaptation for Magnetic Resonance Imaging environment: A systematic review and meta-analysis. Mult. Scler. Relat. Disord. 20, 136–143 (2018).

48. Bragatto, V. S. R., Andrade, L. P. D., Rossi, P. G. & Ansai, J. H. Dual-task during gait between elderly with mild cognitive impairment and Alzheimer: systematic review. Fisioter. Em Mov. 30, 849–857 (2017).

49. Lee, H., Choi, S. & Lee, M. Step Detection Robust against the Dynamics of Smartphones. Sensors 15, 27230–27250 (2015).

50. Shimoyama, I. The Finger-Tapping Test: A Quantitative Analysis. Arch. Neurol. 47, 681 (1990).

51. Van Waes, L., Leijten, M., Mariën, P. & Engelborghs, S. Typing competencies in Alzheimer’s disease: An exploration of copy tasks. Comput. Hum. Behav. 73, 311–319 (2017).

52. McGraw, K. O. & Wong, S. P. Forming inferences about some intraclass correlation coefficients. Psychol. Methods 1, 30–46 (1996).

53. Koo, T. K. & Li, M. Y. A Guideline of Selecting and Reporting Intraclass Correlation Coefficients for Reliability Research. J. Chiropr. Med. 15, 155–163 (2016).

54. Friedman, J., Hastie, T. & Tibshirani, R. Regularization Paths for Generalized Linear Models via Coordinate Descent. J. Stat. Softw. 33, 1–22 (2010).

55. Harrell, J. Regression Modeling Strategies: With Applications to Linear Models, Logistic and Ordinal Regression, and Survival Analysis. (Springer, Cham, 2015). doi:10.1007/978-3-319-19425-7.

57. Wellcome Trust Centre for Neuroimaging. Statistical Parametric Mapping. (2020).

57. Gaser, C. et al. CAT: a computational anatomy toolbox for the analysis of structural MRI data. GigaScience 13, giae049 (2024).

58. The MathWorks Inc. MATLAB 2021b. (2021).

59. Ashburner, J. A fast diffeomorphic image registration algorithm. NeuroImage 38, 95–113 (2007).

60. Brown, T. I., Staresina, B. P. & Wagner, A. D. Noninvasive functional and anatomical imaging of the human medial temporal lobe. Cold Spring Harb. Perspect. Biol. 7, a021840 (2015).

61. Newsome, R. N. et al. Dissociable contributions of thalamic nuclei to recognition memory: novel evidence from a case of medial dorsal thalamic damage. Learn. Mem. 25, 31–44 (2018).

62. Brugnolo, A. et al. Brain Resources: How Semantic Cueing Works in Mild Cognitive Impairment due to Alzheimer’s Disease (MCI-AD). Diagnostics 11, 108 (2021).

63. Swick, D., Ashley, V. & Turken, U. Are the neural correlates of stopping and not going identical? Quantitative meta-analysis of two response inhibition tasks. NeuroImage 56, 1655–1665 (2011).

64. Chang, E. F., Raygor, K. P. & Berger, M. S. Contemporary model of language organization: an overview for neurosurgeons. J. Neurosurg. 122, 250–261 (2015).

65. Starowicz-Filip, A. et al. The role of the cerebellum in the regulation of language functions. Psychiatr. Pol. 51, 661–671 (2017).

66. Oh, A., Duerden, E. G. & Pang, E. W. The role of the insula in speech and language processing. Brain Lang. 135, 96–103 (2014).

67. Baier, B., Müller, N. G. & Dieterich, M. What part of the cerebellum contributes to a visuospatial working memory task?: Cerebellum and Working Memory. Ann. Neurol. 76, 754–757 (2014).

68. Silva, D. et al. Comparison of Four Verbal Memory Tests for the Diagnosis and Predictive Value of Mild Cognitive Impairment. Dement. Geriatr. Cogn. Disord. Extra 2, 120–131 (2012).

69. Vogel, J. W. et al. Subjective cognitive decline and β-amyloid burden predict cognitive change in healthy elderly. Neurology 89, 2002–2009 (2017).

70. Jennings, J. M. & Jacoby, L. L. Improving memory in older adults: Training recollection. Neuropsychol. Rehabil. 13, 417–440 (2003).

71. Gasbarri, A., Pompili, A., Packard, M. G. & Tomaz, C. Habit learning and memory in mammals: Behavioral and neural characteristics. Neurobiol. Learn. Mem. 114, 198–208 (2014).

72. Carpenter, S. K. & DeLosh, E. L. Application of the testing and spacing effects to name learning. Appl. Cogn. Psychol. 19, 619–636 (2005).

73. Chételat, G. et al. Amyloid imaging in cognitively normal individuals, at-risk populations and preclinical Alzheimer’s disease. NeuroImage Clin. 2, 356–365 (2013).

74. Lim, Y. Y. et al. Three-Month Stability of the CogState Brief Battery in Healthy Older Adults, Mild Cognitive Impairment, and Alzheimer’s Disease: Results from the Australian Imaging, Biomarkers, and Lifestyle-Rate of Change Substudy (AIBL-ROCS). Arch. Clin. Neuropsychol. 28, 320–330 (2013).

75. Nelson, P. T. et al. Correlation of Alzheimer disease neuropathologic changes with cognitive status: a review of the literature. J. Neuropathol. Exp. Neurol. 71, 362–381 (2012).

76. Braak, H. & Braak, E. Staging of Alzheimer’s disease-related neurofibrillary changes. Neurobiol. Aging 16, 271–84 (1995).

77. Krumm, S. et al. Cortical thinning of parahippocampal subregions in very early Alzheimer’s disease. Neurobiol. Aging 38, 188–196 (2016).

78. Didic, M. et al. Which memory system is impaired first in Alzheimer’s disease? J. Alzheimers Dis. JAD 27, 11–22 (2011).

79. Kivisaari, S. L., Monsch, A. U. & Taylor, K. I. False positives to confusable objects predict medial temporal lobe atrophy. Hippocampus 23, 832–841 (2013).

80. Bläsi, S. et al. Norms for change in episodic memory as a prerequisite for the diagnosis of Mild Cognitive Impairment (MCI). Neuropsychology 23, 189–200 (2009).

81. O’Bryant, S. E. et al. Staging dementia using Clinical Dementia Rating Scale Sum of Boxes scores: a Texas Alzheimer’s research consortium study. Arch. Neurol. 65, 1091–1095 (2008).

82. Papp, K. V., Rentz, D. M., Orlovsky, I., Sperling, R. A. & Mormino, E. C. Optimizing the preclinical Alzheimer’s cognitive composite with semantic processing: The PACC5. Alzheimers Dement. Transl. Res. Clin. Interv. 3, 668–677 (2017).

