## Supplementary File for "From feasibility to neuroanatomic validity of remote cognitive smartphone assessments in early Alzheimer’s disease"

### 1. Supplementary Material

### 1.1. User Experience Survey and Response Options

A 5-point Likert scale was provided for responses to each question, with response options comprising either “very poor” (1) to “very good” (5) or “I don’t agree at all” (1) to “I completely agree” (5), as appropriate.

1. How would you rate your overall experience with the Digital POC study application and phone?
2. Was the time you needed to complete the smartphone tasks each day acceptable?
3. How difficult was it to plan time to do your smartphone tasks and surveys every day?
4. How burdensome was it for you to complete your smartphone tasks and surveys on a daily basis for the whole duration of the study?
5. How much difficulty did you have putting the study smartphone into the running belt for the 30 Second Walk?
6. How burdensome was it for you to carry the study and your personal smartphones around as much as possible for the duration of the study?
7. If you could remove one component of the Digital POC study application, which one would that be (check only one)? [each component listed]
8. Please rate your agreement with the following statement: The instructions on how to carry out the tasks on the smartphone were clear and easy to understand. What could have been explained better or made easier?
9. Please rate your agreement with the following statement: The materials I received together with the phone were clear and helped me understand how to carry out the tasks. What could have been explained better or made easier?

### 1.2. Study Partner Survey and Response Options

1. Could the study participant do the tasks without assistance? (Yes/No)
2. Did the study participant require any help using the app or doing the tasks in the 30-day period? (Yes/No)
3. What type of support did you give to the study participant? Check all that apply*. (* Multiple responses are possible.)
   a. Reminding the study participant to complete the tasks on the study smartphone.
   b. Giving instructions on how to complete one or more tasks.
   c. Providing physical support to complete the task (e.g. providing aid during the walk task).
   d. Troubleshooting technical issues on the smartphone.
   e. Other.
4. How often did you have to remind the study participant to complete their smartphone tasks?
   a. Never reminded the study participant
   b. Less than once a week
   c. About once a week
   d. 1-2 times a week
   e. 3-5 times a week
   f. 6-7 times a week

#### 1.3. Story Time Data Processing:

Each audio recording was resampled to 16 kHz and its contents classified into intervals of active speech or speech pause, using a speech activity detection algorithm with high temporal resolution (20 ms) [1]. For Narration, relative speech duration was calculated as the cumulative duration of all active speech intervals ≥250 ms, divided by the time between the beginning of the first and the end of the last interval. For Recall, relative speech duration was calculated as the ratio between the cumulative duration of active speech intervals ≥250 ms during Recall, to the corresponding duration in the preceding Narration recording of the same story.

Manual transcriptions of recordings were produced by a professional transcription service [TransPerfect Translations GmbH, Berlin, Germany] with bespoke transcription guidelines to obtain machine-readable, full-verbatim transcriptions (e.g., including incorrect grammar, if applicable), with each utterance on a separate line.

Parts of speech in the transcripts were automatically tagged using the Flair framework [2] using the universal part-of-speech tagging models for English [3] and Spanish [4], respectively. Noun rate was computed as the number of words tagged as nouns, divided by the total number of words.

#### 1.4. Supplementary Tables

Supplementary Table 1. Adherence Overall, by Group, and by AD-DAS Active Task.

| Active Tests | **Median Adherence (%) by Group** | | | | **Overall (%)** |
| --- | --- | --- | --- | --- | --- |
|  | **HC** | **SCDn** | **SCDp** | **eAD** |  |
| Gallery Game Learning | 88.9 | 96.3 | 96.3 | 96.3 | 96.3 |
| Gallery Game Free Recall | 73.3 | 83.3 | 80 | 73.3 | 76.7 |
| Tilt Task | 88.9 | 100 | 100 | 100 | 100 |
| Story Time Narration | 88.9 | 88.9 | 88.9 | 88.9 | 88.9 |
| Story Time Free Recall | 88.9 | 88.9 | 88.9 | 94.4 | 88.9 |
| Object Features | 88.9 | 88.9 | 88.9 | 88.9 | 88.9 |
| Find the Egg | 88.9 | 88.9 | 88.9 | 88.9 | 88.9 |
| Information Processing Speed | 88.9 | 88.9 | 88.9 | 88.9 | 88.9 |
| 30’ Dual Task Walking | 88.9 | 88.9 | 100 | 94.45 | 88.9 |
| Speeded Tapping | 88.9 | 88.9 | 100 | 94.45 | 88.9 |
| Fairytale | 88.9 | 88.9 | 88.9 | 77.8 | 88.9 |
| **Overall** | **89.3** | **96.4** | **96.4** | **96.4** | **96.4** |

AD-DAS=Alzheimer’s Disease Digital Assessment Suite; eAD=early Alzheimer’s Disease; HC=Healthy Controls; SCD=Subjective Cognitive Decline; SCDn=amyloid beta negative SCD; SCDp=amyloid beta positive SCD.

Supplementary Table 2. Known-group comparisons on the AD-DAS active test outcome measures (* for unadjusted p-value ≤ 5x10-2; ** for unadjusted p-value ≤ 5x10-3; and *** for unadjusted p-value ≤ 5x10-4).

| Active Task | Active Task Outcome measure | Group Comparisons | | | | | | | |
| --- | --- | --- | --- | --- | --- | --- | --- | --- | --- |
|  |  | HC vs. eAD | | SCDn vs. eAD | | SCDp vs. eAD | | SCDn vs. SCDp | |
|  |  | DLO | p-value | DLO | p-value | DLO | p-value | DLO | p-value |
| Gallery Game Learning | Total Time | -1.48 | ** | -2.23 | *** | -1.40 | * | -0.84 | 0.08 |
|  | Learning % correct | 0.65 | 0.20 | 0.57 | 0.27 | 0.21 | 0.67 | 0.37 | 0.45 |
| Gallery Game Recognition | Total Time < 24h | -1.04 | *** | -1.83 | *** | -1.03 | *** | -0.80 | ** |
|  | Total Time >24h | -1.96 | *** | -1.78 | *** | -1.44 | *** | -0.35 | 0.16 |
|  | Accuracy <24h | 0.26 | 0.35 | 0.58 | * | 0.05 | 0.86 | 0.53 | 0.05 |
|  | Accuracy >24h | -0.15 | 0.55 | -0.02 | 0.94 | 0.12 | 0.61 | -0.14 | 0.57 |
| Gallery Game Free Recall | Total Time < 24h | -1.11 | *** | -1.84 | *** | -1.24 | *** | -0.60 | * |
|  | Total Time >24h | -2.05 | *** | -1.98 | *** | -1.62 | *** | -0.36 | 0.14 |
|  | % correct <24h | 0.17 | 0.52 | 0.49 | 0.06 | 0.28 | 0.27 | 0.21 | 0.42 |
|  | % correct >24h | -0.23 | 0.36 | -0.15 | 0.55 | 0.07 | 0.76 | -0.22 | 0.37 |
| Tilt Task | Total score | 1.35 | * | 1.50 | ** | 1.38 | * | 0.12 | 0.80 |
| Story Time | Noun rate | 1.54 | * | 1.54 | ** | 1.10 | * | 0.44 | 0.34 |
| Story Time Immediate Free Recall | Ratio of speech duration | 2.08 | *** | 2.22 | *** | 2.29 | *** | -0.06 | 0.90 |
| Story Time Delayed Free Recall | Ratio of speech duration | 1.97 | *** | 1.51 | ** | 1.30 | * | 0.21 | 0.67 |
| Object Features | Mean reaction time for correct answers | -1.43 | ** | -1.58 | ** | -1.56 | ** | -0.02 | 0.97 |
| Find the Egg | Highest level reached | 1.96 | ** | 1.09 | 0.08 | -0.17 | 0.78 | 1.26 | * |
| Information Processing Speed | Number of correct responses | 1.36 | * | 1.64 | ** | 1.45 | * | 0.20 | 0.69 |
| 30’ Dual Task Walking (without gait aid) | 50th %ile step power | 3.28 | *** | 0.43 | 0.43 | 0.68 | 0.20 | -0.25 | 0.60 |
| Speeded Tapping | Mean inter-tap interval - dominant hand | -1.47 | ** | -0.99 | 0.06 | -1.10 | * | 0.11 | 0.83 |
|  | Mean inter-tap interval - non dominant hand | -0.22 | 0.66 | -0.67 | 0.19 | -0.88 | 0.08 | 0.21 | 0.68 |
| Clinical Comparators | | | | | | | | | |
| AIADLQSV | Total Score | 2.17 | *** | 2.34 | *** | 2.03 | *** | 0.31 | 0.51 |
| FCSRT | Free Recall (Encoding) | 3.79 | *** | 2.54 | *** | 2.73 | *** | -0.19 | 0.70 |
| FCSRT | Sensitivity to Cueing (Encoding) | 3.10 | *** | 2.85 | *** | 3.02 | *** | -0.18 | 0.82 |
| FCSRT | Free Recall (30min - Delayed) | 3.49 | *** | 2.55 | *** | 3.67 | *** | -1.12 | * |
| TMT | Time Taken A | -6.95 | *** | -1.71 | ** | -2.21 | *** | 0.51 | 0.29 |
| TMT | Time Taken B - Time Taken A | -4.53 | *** | -1.21 | * | -1.59 | ** | 0.38 | 0.41 |
| BNT | Correct Responses | 2.40 | *** | 0.87 | 0.11 | 2.01 | ** | -1.14 | * |
| VFT | Ratio of Acceptable Words to Words Generated | 2.20 | *** | 1.14 | * | 1.34 | * | -0.20 | 0.70 |
| DSCT | Total Number of Correct Answers | 3.10 | *** | 1.69 | ** | 2.11 | *** | -0.42 | 0.38 |
| T25FWT | Average Time Taken in Seconds | -1.67 | ** | -0.86 | 0.08 | -1.64 | ** | 0.78 | 0.11 |
| TUG | Time in Seconds to Complete Task | -1.55 | ** | -0.58 | 0.25 | -0.75 | 0.13 | 0.17 | 0.73 |
| HADS | Anxiety Total Score | -0.69 | 0.17 | -0.61 | 0.20 | -0.29 | 0.57 | -0.33 | 0.48 |
| HADS | Depression Total Score | -1.04 | 0.08 | -0.66 | 0.22 | -0.46 | 0.39 | -0.20 | 0.70 |
| PSQI | Global Score (derived) | -0.77 | 0.12 | -0.15 | 0.76 | -0.13 | 0.78 | -0.02 | 0.97 |
| NPI | Total Score (derived) | -0.52 | 0.41 | -0.80 | 0.18 | -0.32 | 0.58 | -0.47 | 0.45 |
| UCLA | Total Score | 0.47 | 0.34 | 0.82 | 0.11 | 0.10 | 0.84 | 0.72 | 0.13 |
| SNI | Total Number of People in Social Network | 1.30 | * | -0.32 | 0.57 | 0.39 | 0.45 | -0.70 | 0.17 |

DLO=Differences in Log Odds; 25FWT=25-foot walk test; AIADLQSV=Amsterdam Instrumental Activities of Daily Living Questionnaire Short Version; BNT=Boston Naming Test, DSCT=Digit‑Symbol Coding Test; dom=dominant; eAD=early Alzheimer’s Disease; FCSRT=Free and Cued Selective Reminding Test; HADS=Hospital Anxiety and Depression Scale; HC=Healthy Controls; LOR=log odds ratio; MMSE=Mini-Mental State Examination; nondom=non-dominant; NPI=Neuropsychiatric Inventory; PSQI=Pittsburgh Sleep Quality Index; SCD=Subjective Cognitive Decline; SCDn=amyloid beta negative SCD; SCDp=amyloid beta positive SCD; TMT=Trail-Making Test; TUG=Timed Up and Go;UCLA=University of California, Los Angeles; VFT=Verbal Fluency Task. Dashes in convergent validity signifies no or negligible variance in the resulting clinical comparator, and hence the model did not converge or no informed statistics could be performed

Supplementary Table 3. Results of voxel-based morphometry analyses of anatomic MRI scans correlating AD-DAS and clinical neuropsychological outcome measures with estimates of gray matter integrity across the entire brain. All correlations are positive unless otherwise noted.

| **Digital measures** | | | | | | | | | | **Clinical measures** | | | | | | | | | |
| --- | --- | --- | --- | --- | --- | --- | --- | --- | --- | --- | --- | --- | --- | --- | --- | --- | --- | --- | --- |
| **AD-DAS task** | **Outcome measure** | **Cluster-Level** | | **Voxel-Level** | | **MNI Coordinates of Peak Voxel** | | | **Location of Peak Voxel** | **Clinical task** | **Outcome measure** | **Cluster-level** | | **Voxel-level** | | **MNI Coordinates of Peak Voxel** | | | **Location of Peak Voxel** |
|  |  | **FWE p‑value** | **Extent** | ***t*‑score** | **FWE p‑value** | ***x*** | ***y*** | ***z*** |  |  |  | **FWE p‑value** | **Extent** | ***t*‑score** | **FWE *p*‑value** | ***x*** | ***y*** | ***z*** |  |
| Gallery Game Learning | Total time (negative correlation) | <0.0001 | 2103 | 5.43 | 0.01 | -26 | 14 | -32 | Left superior temporal pole | FCSRT | Free recall (encoding) | <0.0001 | 4447 | 6.18 | 0.001 | -32 | -20 | -12 | Left hippocampus |
|  |  |  |  | 5.43 | 0.01 | -23 | 6 | -35 |  |  |  |  |  | 6 | 0.001 | -24 | -36 | 2 |  |
|  |  |  |  | 5.33 | 0.015 | -32 | 15 | -26 |  |  |  |  |  | 5.41 | 0.012 | -18 | -14 | -14 |  |
|  |  | <0.0001 | 1469 | 5.38 | 0.013 | 38 | -83 | 8 | Right middle occipital lobe |  |  | <0.0001 | 4986 | 5.85 | 0.002 | 35 | -24 | -17 | Right hippocampus |
|  |  |  |  | 5.16 | 0.028 | 47 | -87 | -8 |  |  |  |  |  | 5.57 | 0.007 | 24 | -33 | -3 |  |
|  |  |  |  | 4.98 | 0.054 | 33 | -93 | 3 |  |  |  |  |  | 5.08 | 0.041 | 26 | -11 | -18 |  |
|  |  | 0.032 | 493 | 4.51 | 0.248 | 20 | 9 | -35 | Right perirhinal cortex |  |  | 0 | 1066 | 5.38 | 0.014 | 59 | -29 | -21 | Right inferior temporal lobe |
|  |  |  |  | 4.04 | 0.717 | 26 | 5 | -32 |  |  |  |  |  | 5.2 | 0.027 | 66 | -12 | -24 |  |
|  |  |  |  | 3.93 | 0.825 | 27 | 17 | -29 |  |  |  |  |  | 4.51 | 0.262 | 65 | -42 | -15 |  |
|  |  |  | | | | | | | |  |  | 0.003 | 769 | 4.62 | 0.193 | -45 | 38 | -2 | Left inferior frontal gyrus (triangular) |
|  |  |  |  |  |  |  |  |  |  |  |  |  |  | 4.32 | 0.433 | -36 | 29 | 3 |  |
|  |  |  |  |  |  |  |  |  |  |  |  |  |  | 4.15 | 0.614 | -44 | 47 | 0 |  |
|  |  |  |  |  |  |  |  |  |  |  |  | 0.01 | 622 | 4.24 | 0.522 | -60 | -33 | 2 | Left middle temporal lobe |
|  |  |  |  |  |  |  |  |  |  |  |  |  |  | 4.14 | 0.626 | -63 | -45 | 6 |  |
|  |  |  |  |  |  |  |  |  |  |  |  |  |  | 3.68 | 0.977 | -63 | -23 | 0 |  |
|  |  |  |  |  |  |  |  |  |  |  |  | 0.012 | 604 | 3.82 | 0.92 | -18 | -14 | 11 | Left thalamus |
|  |  |  |  |  |  |  |  |  |  |  |  |  |  | 3.69 | 0.976 | 0 | -6 | 9 |  |
|  |  |  |  |  |  |  |  |  |  |  |  |  |  | 3.29 | 1 | -6 | -24 | 12 |  |
| Gallery Game Recognition < 24h | Hit rate | 0.002 | 881 | 4.72 | 0.133 | -6 | -20 | 0 | Left thalamus | FCSRT | Sensitivity to cueing | <0.0001 | 1786 | 7.68 | <0.0001 | -18 | 17 | -20 | Left medial orbitofrontal cortex |
|  |  |  |  | 4.62 | 0.184 | -5 | -23 | 9 |  |  |  |  |  | 5.16 | 0.029 | -42 | 40.5 | -12 |  |
|  |  |  |  | 4.09 | 0.673 | -18 | -20 | 8 |  |  |  |  |  | 4.6 | 0.193 | -18 | 36 | -17 |  |
|  |  |  | | | | | | | |  |  | 0 | 1314 | 6.83 | <0.0001 | 18 | 17 | -18 | Right gyrus rectus next to superior orbitofrontal lobe |
|  |  |  |  |  |  |  |  |  |  |  |  |  |  | 5.39 | 0.012 | 30 | 11 | -21 |  |
|  |  |  |  |  |  |  |  |  |  |  |  |  |  | 3.89 | 0.859 | 9 | 36 | -14 |  |
|  |  |  |  |  |  |  |  |  |  |  |  | <0.0001 | 1945 | 6.51 | 0 | -57 | -9 | -26 | Left middle temporal lobe |
|  |  |  |  |  |  |  |  |  |  |  |  |  |  | 6.02 | 0.001 | -47 | -6 | -41 |  |
|  |  |  |  |  |  |  |  |  |  |  |  |  |  | 5.16 | 0.029 | -56 | -24 | -24 |  |
|  |  |  |  |  |  |  |  |  |  |  |  | 0.001 | 965 | 6.12 | 0.001 | 56 | -24 | -27 | Right inferior temporal lobe |
|  |  |  |  |  |  |  |  |  |  |  |  |  |  | 5.83 | 0.002 | 53 | -30 | -21 |  |
|  |  |  |  |  |  |  |  |  |  |  |  |  |  | 3.87 | 0.876 | 60 | -21 | -21 |  |
| Gallery Game Free Recall < 24h | Total time (negative correlation) | 0.003 | 806 | 4.12 | 0.641 | -23 | 5 | -36 | Left perirhinal cortex | FCSRT | Delayed free recall | 0 | 1266 | 5.82 | 0.002 | -24 | -36 | 1.5 | Left hippocampus |
|  |  |  |  |  |  |  |  |  |  |  |  |  |  | 4.69 | 0.156 | -29 | -30 | -9 |  |
|  |  |  |  | 4.03 | 0.738 | -32 | 15 | -24 |  |  |  |  |  | 4.6 | 0.201 | -15 | -12 | -15 |  |
|  |  |  |  | 3.96 | 0.809 | -44 | 20 | -15 |  |  |  | <0.0001 | 2018 | 4.78 | 0.113 | 24 | -33 | -3 | Right hippocampus |
|  |  |  | | | | | | | |  |  |  |  | 4.48 | 0.282 | 38 | -23 | -18 |  |
|  |  |  |  |  |  |  |  |  |  |  |  |  |  | 4.34 | 0.411 | 29 | 17 | -29 |  |
|  |  |  |  |  |  |  |  |  |  |  |  | 0.001 | 988 | 4.47 | 0.29 | -36 | 6 | -33 | Left middle temporal lobe |
|  |  |  |  |  |  |  |  |  |  |  |  |  |  | 4.32 | 0.427 | -26 | 9 | -41 |  |
|  |  |  |  |  |  |  |  |  |  |  |  |  |  | 4.18 | 0.584 | -26 | 14 | -32 |  |
| Tilt Task | % correct in response inhibition trials (normalized by non-response inhibition trials) | <0.0001 | 1518 | 4.98 | 0.071 | 42 | 3 | 2 | Right insula | TMT | Time B-A | *No supra-threshold clusters* | | | | | | | |
|  |  |  |  | 4.58 | 0.248 | 41 | -12 | 8 |  |  |  |  |  |  |  |  |  |  |  |
|  |  |  |  | 4.27 | 0.54 | 35 | -14 | 14 |  |  |  |  |  |  |  |  |  |  |  |
|  |  | <0.0001 | 1356 | 4.5 | 0.309 | -38 | -2 | -39 | Left inferior temporal lobe |  |  |  |  |  |  |  |  |  |  |
|  |  |  |  | 4.47 | 0.341 | -27 | 11 | -39 |  |  |  |  |  |  |  |  |  |  |  |
|  |  |  |  | 4.42 | 0.381 | -42 | 8 | -42 |  |  |  |  |  |  |  |  |  |  |  |
|  |  | <0.001 | 1039 | 5.21 | 0.033 | -33 | 18 | 3 | Left insula |  |  |  |  |  |  |  |  |  |  |
|  |  |  |  | 4.56 | 0.267 | -33 | 20 | -8 |  |  |  |  |  |  |  |  |  |  |  |
|  |  |  |  | 4.24 | 0.567 | -32 | 33 | -3 |  |  |  |  |  |  |  |  |  |  |  |
|  |  | <0.001 | 943 | 5.24 | 0.029 | -35 | -20 | -12 | Left hippocampus |  |  |  |  |  |  |  |  |  |  |
|  |  |  |  | 4.89 | 0.097 | -23 | -15 | -12 |  |  |  |  |  |  |  |  |  |  |  |
|  |  |  |  | 4.44 | 0.365 | -26 | -33 | 0 |  |  |  |  |  |  |  |  |  |  |  |
|  |  | 0.004 | 726 | 5.03 | 0.0605 | 24 | -20 | -10 | Right hippocampus |  |  |  |  |  |  |  |  |  |  |
|  |  |  |  | 4.38 | 0.423 | 36 | -24 | -17 |  |  |  |  |  |  |  |  |  |  |  |
|  |  |  |  | 3.93 | 0.873 | 30 | -8 | -12 |  |  |  |  |  |  |  |  |  |  |  |
|  |  | 0.012 | 579 | 4.27 | 0.535 | 27 | 3 | -44 | Right fusiform gyrus |  |  |  |  |  |  |  |  |  |  |
|  |  |  |  | 4.22 | 0.593 | 33 | 11 | -38 |  |  |  |  |  |  |  |  |  |  |  |
|  |  |  |  | 3.78 | 0.959 | 39 | 8 | -42 |  |  |  |  |  |  |  |  |  |  |  |
|  |  | 0.023 | 500 | 5.32 | 0.022 | 20 | -95 | -6 | Right calcarine sulcus |  |  |  |  |  |  |  |  |  |  |
|  |  |  |  | 3.58 | 0.996 | 5 | -95 | -5 |  |  |  |  |  |  |  |  |  |  |  |
| Story Time Narration | Speech duration divided by pause duration at ST Narration; negative correlation) | 0.015 | 570 | 4.1 | 0.669 | -26 | -77 | -24 | Left cerebellum | BNT | Number correct | *No supra-threshold clusters* | | | | | | | |
|  |  |  |  | 4.01 | 0.762 | -20 | -66 | -35 |  |  |  |  |  |  |  |  |  |  |  |
|  |  | 0.03 | 472 | 4.05 | 0.728 | -57 | -62 | 24 | Left angular gyrus | VFT | Number correct | *No supra-threshold clusters* | | | | | | | |
|  |  |  |  | 3.97 | 0.8 | -56 | -59 | 15 |  |  |  |  |  |  |  |  |  |  |  |
|  |  |  |  | 3.95 | 0.819 | -62 | -51 | 8 |  |  |  |  |  |  |  |  |  |  |  |
| Story Time Immediate Free Recall <24h | Speech duration at ST Immediate Free Recall divided by speech duration at ST Narration | <0.0001 | 2448 | 5.48 | 0.01 | -38 | -22 | -16 | Left fusiform gyrus | *See above results for* FCSRT Free Recall (encoding) *and* FCSRT Delayed free recall | | | | | | | | | |
|  |  |  |  | 4.89 | 0.081 | -21 | -18 | -14 |  |  |  |  |  |  |  |  |  |  |  |
|  |  |  |  | 4.44 | 0.327 | -36 | -15 | -32 |  |  |  |  |  |  |  |  |  |  |  |
|  |  | 0.0001 | 1297 | 5.47 | 0.01 | -57 | -18 | -2 | Left middle temporal lobe |  |  |  |  |  |  |  |  |  |  |
|  |  |  |  | 5.27 | 0.021 | -63 | -12 | -9 |  |  |  |  |  |  |  |  |  |  |  |
|  |  |  |  | 4.8 | 0.109 | -48 | -8 | -14 |  |  |  |  |  |  |  |  |  |  |  |
|  |  | <0.0001 | 1841 | 5.15 | 0.032 | 23 | -20 | -11 | Right hippocampus |  |  |  |  |  |  |  |  |  |  |
|  |  |  |  | 4.91 | 0.075 | 36 | -26 | -17 |  |  |  |  |  |  |  |  |  |  |  |
|  |  |  |  | 4.89 | 0.081 | 21 | -20 | -23 |  |  |  |  |  |  |  |  |  |  |  |
|  |  | 0.019 | 535 | 4.64 | 0.183 | 44 | 2 | 0 | Right insula |  |  |  |  |  |  |  |  |  |  |
|  |  |  |  | 3.95 | 0.827 | 44 | -6 | 12 |  |  |  |  |  |  |  |  |  |  |  |
|  |  |  |  | 3.45 | 0.999 | 39 | 6 | 17 |  |  |  |  |  |  |  |  |  |  |  |
|  |  | 0.01 | 614 | 4.6 | 0.204 | 68 | -23 | -26 | Right inferior temporal lobe |  |  |  |  |  |  |  |  |  |  |
|  |  |  |  | 4.26 | 0.496 | 63 | -9 | -30 |  |  |  |  |  |  |  |  |  |  |  |
|  |  |  |  | 3.84 | 0.911 | 60 | -17 | -24 |  |  |  |  |  |  |  |  |  |  |  |
|  |  | 0.0001 | 1933 | 4.46 | 0.302 | -3 | -62 | 6 | Left lingual gyrus |  |  |  |  |  |  |  |  |  |  |
|  |  |  |  | 4.4 | 0.355 | -6 | -47 | 24 |  |  |  |  |  |  |  |  |  |  |  |
|  |  |  |  | 4.31 | 0.451 | -3 | -36 | 23 |  |  |  |  |  |  |  |  |  |  |  |
|  |  | 0.039 | 447 | 4.38 | 0.38 | 23 | -54 | -12 | Right fusiform |  |  |  |  |  |  |  |  |  |  |
|  |  | 0.011 | 601 | 4.24 | 0.518 | 45 | -14 | 2 | Right superior temporal lobe |  |  |  |  |  |  |  |  |  |  |
|  |  |  |  | 4.11 | 0.668 | 44 | -35 | 18 |  |  |  |  |  |  |  |  |  |  |  |
|  |  |  |  | 3.99 | 0.795 | 50 | -21 | 2 |  |  |  |  |  |  |  |  |  |  |  |
| Find the Egg | Highest level reached | 0.045 | 429 | 4.21 | 0.577 | -29 | -56 | -47 | Left cerebellum 8 | *No clinical comparator* | | | | | | | | | |
|  |  |  |  | 4.19 | 0.591 | -33 | -45 | -50 |  |  |  |  |  |  |  |  |  |  |  |
|  |  |  |  | 3.72 | 0.97 | -44 | -50 | -57 |  |  |  |  |  |  |  |  |  |  |  |
| IPS | Number correct digit-symbol matching in 90 seconds | <0.0001 | 2251 | 5.83 | 0.002 | 57 | -14 | 15 | Right insula | DSCT | Total number correct | *No supra-threshold clusters* | | | | | | | |
|  |  |  |  | 4.6 | 0.206 | 45 | -22 | 14 |  |  |  |  |  |  |  |  |  |  |  |
|  |  |  |  | 4.23 | 0.536 | 48 | -6 | 0 |  |  |  |  |  |  |  |  |  |  |  |
|  |  | 0.006 | 660 | 5.21 | 0.026 | 14 | -64 | -8 | Right lingual gyrus |  |  |  |  |  |  |  |  |  |  |
|  |  |  |  | 4.31 | 0.451 | 24 | -56 | -4 |  |  |  |  |  |  |  |  |  |  |  |
|  |  |  |  | 4.24 | 0.52 | 16 | -68 | 2 |  |  |  |  |  |  |  |  |  |  |  |
|  |  | 0.021 | 517 | 4.83 | 0.1 | -36 | -24 | -16 | Left hippocampus |  |  |  |  |  |  |  |  |  |  |
|  |  |  |  | 3.81 | 0.93 | -34 | -30 | -26 |  |  |  |  |  |  |  |  |  |  |  |
|  |  |  |  | 3.63 | 0.986 | -27 | -38 | -16 |  |  |  |  |  |  |  |  |  |  |  |
|  |  | 0.017 | 543 | 4.73 | 0.139 | -10 | -57 | 26 | Left precuneus |  |  |  |  |  |  |  |  |  |  |
|  |  |  |  | 4.38 | 0.38 | -3 | -56 | 34 |  |  |  |  |  |  |  |  |  |  |  |
|  |  |  |  | 4.12 | 0.662 | -8 | -64 | 32 |  |  |  |  |  |  |  |  |  |  |  |
|  |  | 0.043 | 431 | 4.69 | 0.154 | 27 | -38 | -15 | Right fusiform gyrus |  |  |  |  |  |  |  |  |  |  |
|  |  |  |  | 3.38 | 0.999 | 33 | -22 | -20 |  |  |  |  |  |  |  |  |  |  |  |

BNT = Boston Naming Test; DSCT= Digital Symbol Coding Test; FCSRT= Free and Cued Selective Reminding Test; FWE = family-wise error; IPS = Information Processing Speed; MNI = Montreal Neurological Institute; TMT = Trail Making Test; VFT = Verbal Fluency Test.

#### 1.5. Supplementary Figures

Supplementary Figure 1. Convergent validity scatterplot for each selected feature and the corresponding clinical comparator. Coloured by participant group; Green: Healthy Controls (HC), Turquoise: amyloid beta negative SCD (SCDn), Purple: amyloid beta positive SCD (SCDp), Orange: early Alzheimer’s Disease (eAD).

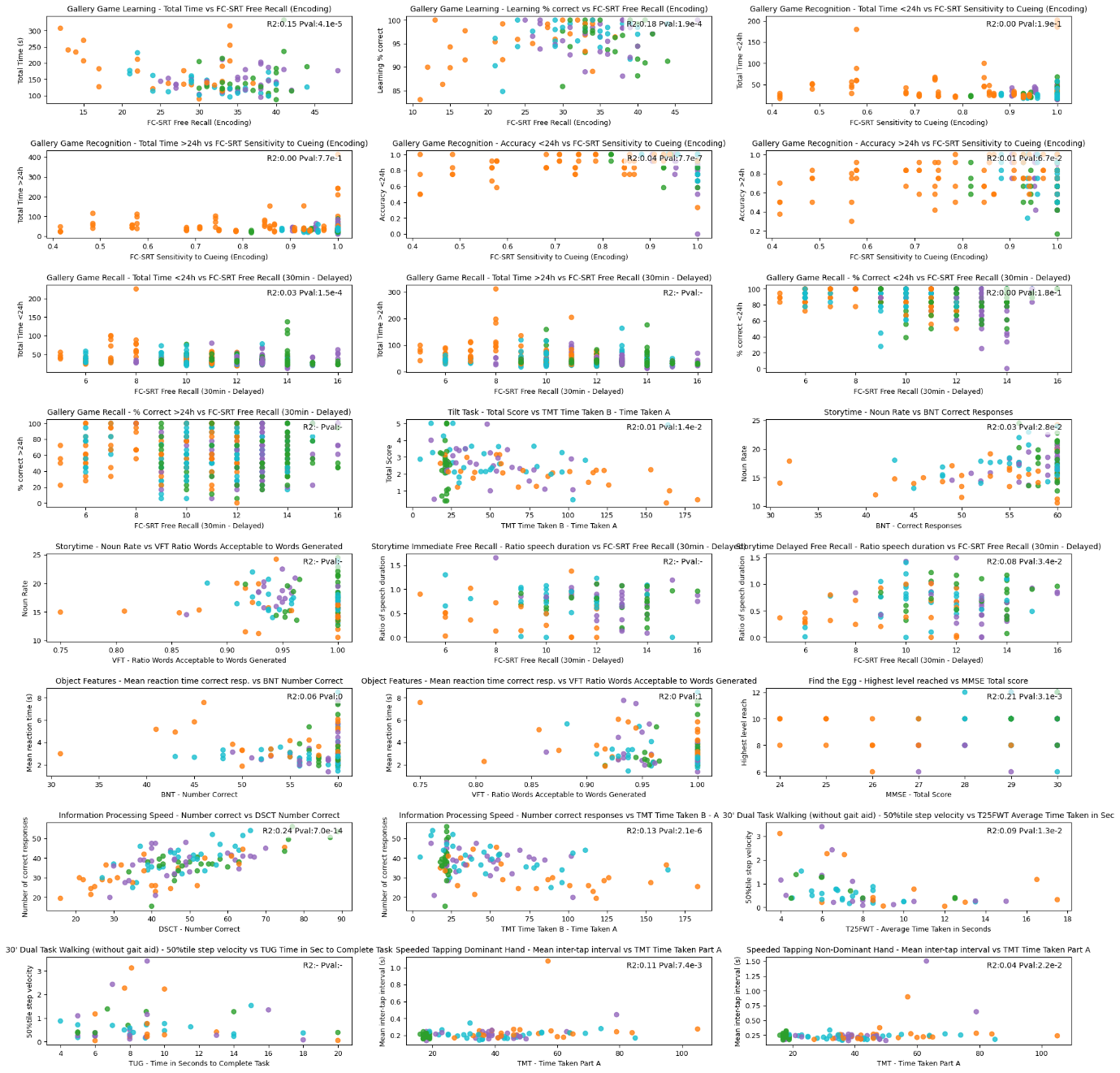

Supplementary Figure 2. Known-groups validity boxplot for each selected feature. Coloured by participant group; Green: Healthy Controls (HC), Turquoise: amyloid beta negative SCD (SCDn), Purple: amyloid beta positive SCD (SCDp), Orange: early Alzheimer’s Disease (eAD).

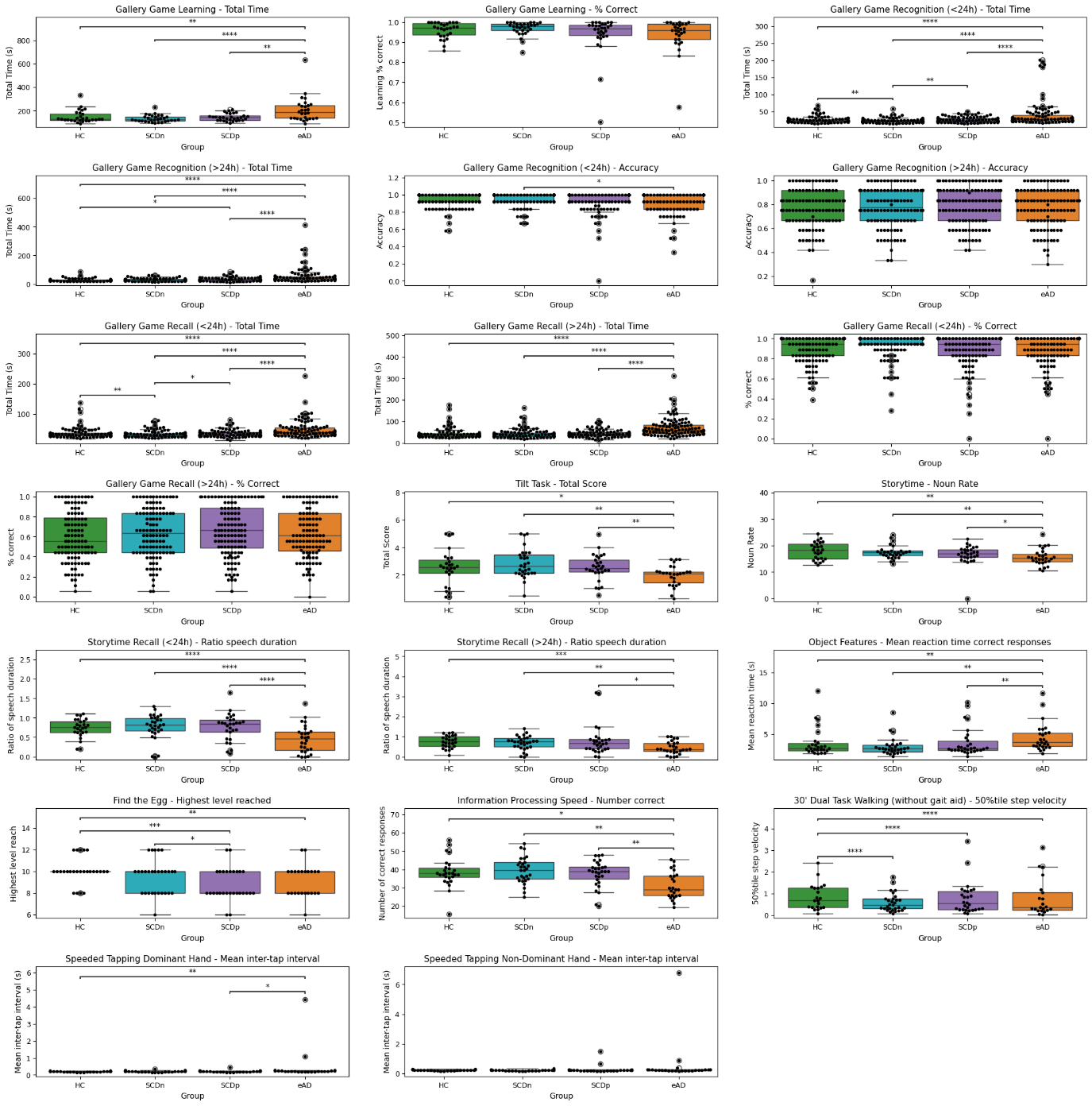

#### 1.6. References

[1]	WebRTC Contributors. Py-Webrtcvad 2021.

[2]	Akbik A, Bergmann T, Blythe D, Rasul K, Schweter S, Vollgraf R. FLAIR: An Easy-to-Use Framework for State-of-the-Art NLP. In: Ammar W, Louis A, Mostafazadeh N, editors. Proceedings of the 2019 Conference of the North American Chapter of the Association for Computational Linguistics (Demonstrations), Minneapolis, Minnesota: Association for Computational Linguistics; 2019, p. 54–9. https://doi.org/10.18653/v1/N19-4010.

[3]	Akbik A, Blythe D, Vollgraf R. Contextual String Embeddings for Sequence Labeling. In: Bender EM, Derczynski L, Isabelle P, editors. Proceedings of the 27th International Conference on Computational Linguistics, Santa Fe, New Mexico, USA: Association for Computational Linguistics; 2018, p. 1638–49.

[4]	Taulé M, Martí MA, Recasens M. AnCora: Multilevel Annotated Corpora for Catalan and Spanish. In: Calzolari N, Choukri K, Maegaard B, Mariani J, Odijk J, Piperidis S, et al., editors. Proceedings of the Sixth International Conference on Language Resources and Evaluation (LREC’08), Marrakech, Morocco: European Language Resources Association (ELRA); 2008.
